# Observations of respiratory syncytial virus (RSV) nucleic-acids in wastewater solids across the United States in the 2022-2023 season: Relationships with RSV infection positivity and hospitalization rates

**DOI:** 10.1101/2023.11.16.23298599

**Authors:** Alessandro Zulli, Meri R.J. Varkila, Julie Parsonnet, Marlene K. Wolfe, Alexandria B. Boehm

**Affiliations:** Department of Civil and Environmental Engineering, Stanford University, 473 Via Ortega, Stanford, California, 94305; Division of Infectious Diseases and Geographic Medicine, Department of Medicine, Stanford University, 300 Pasteur Drive, Stanford, California, 94305; Department of Epidemiology and Population Health, Stanford University, 300 Pasteur Drive, Stanford, California, 94305; Gangarosa Department of Environmental Health, Rollins School of Public Health, Emory University, 1518 Clifton Rd, Atlanta, GA, USA, 30322

**Author notes:** These individuals contributed equally to this work (Alessandro Zulli, Meri R.J. Varkila, Marlene K. Wolfe).

## Abstract

Respiratory syncytial virus (RSV) is a leading cause of respiratory illness and hospitalization but surveillance detects only a minority of cases. Wastewater surveillance could determine onset and extent of RSV circulation in the absence of sensitive case detection, but to date, studies of RSV in wastewater have been few. We measured RSV genomic RNA concentrations in wastewater solids from 176 US sites that provided samples at least three times per week during the 2022-2023 RSV season. Concentrations ranged from undetectable to 10^7^ copies per gram dry weight (median = 10^3^ cp/g). Wastewater RSV RNA concentration aggregated at state and national levels correlated with case positivity and hospitalization rates. Onset of RSV season was assessed for 14 states that collected data prior to the start of the RSV season. Wastewater and clinical surveillance identified onset of RSV during the same week in 3 states, whereas in 3 states, wastewater onset preceded clinical onset, and in 7 states, wastewater onset occurred after clinical onset. Wastewater concentrations generally peaked in the same week as hospitalization rates, but after case positivity rates peaked. Differences in onset and peaks in the wastewater versus clinical data may reflect inherent differences in the surveillance approaches.

## Introduction

Respiratory syncytial virus (RSV) is a leading cause of pediatric pneumonia and bronchiolitis and is responsible for over 100,000 pediatric deaths worldwide each year.^1–3^ In the US, RSV annually accounts for up to 80,000 hospitalizations and over 520,000 emergency department visits in children under 5.^4–6^ RSV infects 97% of all children by the age of 2, making it a ubiquitous cause of acute respiratory tract infection in children, and a large driver of morbidity and mortality within this cohort.^1,7,8^ RSV is also a concern in the aging population, responsible for up to 10,000 recorded yearly deaths in the cohort aged 65 and older.^5,9^

RSV transmission typically follows a predictable seasonal pattern, with peaks in the Northern Hemisphere occurring in the late fall and extending through spring.^10^ Pinpointing the onset and severity of the RSV season could enhance efficient allocation of resources to where and when they are needed and signal public health authorities to campaign immunization. In the U.S. RSV surveillance is conducted through several networks that track temporal and geographic trends in clinical illness, hospitalizations and demographic characteristics of patients.^11^ The determination of epidemic onset is currently tied to laboratory-based testing of clinical cases – a metric based on voluntary reporting that does not capture the full magnitude of RSV circulation in the community and is prone to reporting time lags.^12^ As a result the annual onset of RSV transmission at the community level may only be recognized weeks after it has begun.

During the COVID-19 pandemic, many countries, including the US, observed drastic reductions in RSV infections the winter of 2020-21 followed by unseasonal RSV activity in the summer of 2021 and atypically early onset of RSV circulation with more severe clinical presentation of disease during the winter of 2022-2023.^10–13^ During this period, new immunizations became available and include a vaccine for use during pregnancy,a long-acting monoclonal antibody (nirsevimab) recommended for immunization of infants 8 months old and younger, and a new vaccine available for people 60 and older. These recent changes in RSV circulation and tools for response make determination of the onset of annual RSV epidemics an especially important, yet difficult, task.

Wastewater provides a composite biological sample for a community that can be used to monitor local occurrence of infectious diseases. Previous studies have demonstrated that wastewater concentrations of RSV RNA correlate with disease occurrence within the communities contributing to wastewater^14–21^, however these studies have been conducted at small geographic scales. We have previously demonstrated that wastewater-based measurements of influenza can be related to traditional surveillance metrics across the U.S.^22^ In this study, we use data from across the US to investigate how RSV RNA concentrations in the solids from wastewater relate to traditional disease metrics during the 2022-2023 RSV season. We also test the feasibility of using wastewater-based estimations to determine the onset of RSV season.

## Methods

We conducted a retrospective observational time series analysis using data on RSV RNA concentrations in wastewater collected as part of this study along with publicly available clinical and laboratory data on cases and hospital admissions. Data between 1 January 2022 to 31 July 2023 were used. This study analyzed publicly available clinical data that does not contain protected health information and was therefore exempt from ethics review and the need for informed consent as per Common Rule 45 CFR46.102.

### Clinical surveillance data

The CDC National Respiratory and Enteric Virus Surveillance System (NREVSS) is a laboratory-based surveillance system that records voluntarily reported data on laboratory testing, including confirmed RSV infections, on a weekly basis from participating laboratories nationwide.^23^ The number of PCR tests performed and the proportion of positive PCR results for RSV were extracted by state and at a national level directly from the website and are available for each morbidity and mortality weekly report (MMWR) week as 3-week moving averages; this proportion is referred to as “positivity rate”. We did not utilize results from antigen testing. We obtained RSV hospitalization rates from the CDC Respiratory Syncytial Virus Hospitalization Surveillance Network (RSV-Net), a platform that conducts surveillance for laboratory-confirmed hospital admissions associated with RSV in 12 states.^24^ Hospitalization rate data are available on a weekly basis.

### Wastewater data: sample collection

Between 1 January 2022 and 31 July 2023, wastewater samples (either 24-hour composited influent of grab samples from the primary clarifier, Table S1) were collected by wastewater treatment plant (WWTP) staff using sterile containers. Samples were typically obtained three times per week and shipped overnight to our laboratory at 4°C where they were processed immediately. During the time between sample collection and transport (0-3 days), we expect minimal losses of the RNA target based on results of viral RNA persistence studies.^25^ Samples were collected from WWTPs in a total of 34 states over the course of the study period and a total of 176 distinct WWTPs. The date when sample collection commenced varied by WWTP, depending on when it enrolled in the study (Table S1). A total of 22,809 samples were collected and analyzed as part of this study.

Previous work has indicated that RSV RNA in wastewater strongly adsorbs to wastewater solids, so in this study, we make measurements in the solid phase of wastewater.^26^ Details of the isolation of solids from the samples are provided in other peer-reviewed publications.^27^ In short, samples were centrifuged to dewater the solids and an aliquot was used for nucleic-acid extractions, and another used to determine the dry weight of the solids.

### Wastewater data: Nucleic-acid extraction

The methods for nucleic acid extraction and purification are provided on protocols.io.^28^ In brief, dewatered solids were suspended to DNA/RNA shield (Zymo, Irvine, CA) containing bovine coronavirus (BCoV, an extraction control). The concentration of solids in the DNA/RNA shield approximately 75 mg/ml; we found that there is limited inhibition of downstream analytical measurements using this concentration of solids.^27^ The suspension was homogenized and then centrifuged, and nucleic acids were extracted from 6-10 aliquots of the supernatant using a commercially available kit. The resultant extracts were subjected to inhibitor removal using a commercial kit. 300 µl entered each extraction resulting in 50 µl of nucleic-acid extract for each of the replicates. The number of replicates varied by WWTP as indicated in Figure S1.

### Wastewater data: Analytical methods

Concentrations of RSV, pepper mild mottle virus (PMMoV), and BCoV RNA in each extract were measured using droplet digital RT-PCR. Primers for RSV were previously developed and detect both RSV A and RSV B.^15^ PMMoV is used as a whole process endogenous control, and a fecal strength control; it is a highly abundant plant virus present in wastewater globally.^29,30^ BCoV served as an extraction control. Primers and probes for all the assays were purchased from IDT (Coralville, Iowa) and are presented in Table S2.

PMMoV and BCoV were measured using a duplex assay previously described using a 1:100 dilution of the nucleic-acid extracts as template.^27^ Two replicate wells were run for the PMMoV/BCoV assays, with 10 replicate wells run for a small subset of the WWTPs (Figure S1). Each of the 6-10 replicate nucleic-acid extractions obtained from each sample were run neat as template in their own RT-PCR well to measure RSV RNA concentrations. The RSV assay was run in a multiplex reaction with assays for other targets; the precise other targets included varied over the course of the prospective study as public health needs and interests changed with emerging outbreaks and science to support the use of wastewater for a broad range of infectious disease targets. The precise assays that the RSV assay was multiplexed with are presented in Figure S1. Additional details of the RT-PCR assays (cycling conditions, thresholding) are presented in the SI.^27^ Extraction and PCR positive and negative controls were run on each 96-well plate, as described elsewhere.^27^

Concentrations of the targets are presented as copies per gram dry weight. For a sample to be scored as a positive, there had to be at least 3 positive droplets. The lower detection limit for RSV is approximately 1000 copies/g dry weight. Errors are reported as standard deviations on the measurements as obtained from the instrument software as the “total error”. Wastewater data are publicly available through the Stanford Digital Repository (https://purl.stanford.edu/hd388xb1982).

### Spatial-aggregation of wastewater data

We aggregated data across states and across the nation using the following methods. First, state-aggregated population weighted averages were calculated for each state with at least one WWTP participating in the project using the following equation:

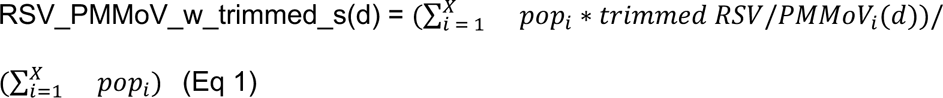

where RSV_PMMoV_w_trimmed_s(d) is the state-aggregated population weighted average of RSV normalized by PMMoV for state s on day d, pop_i_ is the population served by WWTP i of X total in the state, and *trimmed RSV/PMMoV_i_(d)* is the 5-adjacent sample centered, trimmed average of RSV/PMMoV for treatment plant i on day d. Prior to calculating RSV_PMMoV_w_trimmed_s(d), non-detect values were set to 500 cp/g (approximately half the limit of detection) divided by the PMMoV value for the plant that day d, and values for *trimmed RSV/PMMoV_i_(d)* days without data were interpolated using a linear interpolation from adjacent data points. We also calculated RSV_w_trimmed_s(d) which is analogous to RSV_PMMoV_w_trimmed_s(d) but uses *RSV_w_trimmed* rather than trimmed RSV/PMMoV_i_ in Eq. 2.

We aggregated data across the nation using the following equation:

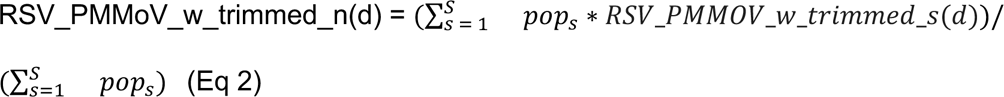

where RSV_PMMoV_w_trimmed_n(d) is the nationally aggregated average of wastewater concentrations of RSV/PMMoV as a function of day, pop_s_ is the population of state s of S states included in the study, and RSV_PMMOV_w_trimmed_s(d) is defined above in Eq 1. We also calculated RSV_w_trimmed_n(d) which is analogous to RSV_PMMoV_w_trimmed_n(d) but uses *RSV_w_trimmed_s* rather than *RSV_PMMOV_w_trimmed_s* in Eq. 2.

In cases where data from a specific WWTP had significant gaps between observations (21 or more days of no observation), the data from that WWTP were not included in the state level aggregation for those periods, but reincluded once observations resumed.

### Data analysis: Correlations

To test associations between wastewater and clinical measures, we calculated spearman’s rho correlations between wastewater concentrations and positivity rates and between wastewater concentrations and hospitalizations for states where both datasets were available. A non-parametric method was chosen as the data tend to not be normally distributed. As clinical data are available on a weekly basis, we used the weekly median RSV_PMMoV_w_trimmed_s(d) or RSV_PMMoV_w_trimmed_n(d) depending on whether the correlations were done at the state or national level, respectively. Weeks were defined as by the CDC Morbidity and Mortality Weekly Report (MMWR). We conducted a total of 44 correlations and to account for multiple correlations used a p value of 0.001 to identify statistically significant correlations for alpha < 0.05 (0.05/44 = 0.001).

### Data analysis: RSV season characteristics

The onset and offset of RSV season were determined using both clinical metrics and wastewater. States for which wastewater data did not exist prior to the clinical onset date of the 2022-2023 RSV season, as determined by positivity rate data, were excluded from this analysis.

Epidemic onset and offset were determined at state-, and national-level based on the first and last of two consecutive weeks when the weekly percentage of positive PCR tests for RSV was greater than or equal to 3%.^31^ From here on, these weeks will be referred to as the clinical onset and offset, and defined by the last day of the week. The duration of the epidemic was defined as the number of weeks between onset and offset.

We recorded the median of RSV_PMMoV_w_trimmed_s during the weeks of clinical onset and offset, for each state. Similarly, the median of RSV_PMMoV_w_trimmed_n was noted for the week during which clinical onset and offset was identified for the country.

The onset and offset of RSV wastewater events were determined using the following approach. First, we used the criteria described by Boehm et al. to identify onset and offset for each individual WWTP as follows.^21^ For each individual WWTP, the wastewater onset date was identified as the date on which all samples in a 14-d look back period had measured concentrations of RSV RNA that exceeded or were equal to 2000 cp/g dry weight. The wastewater offset date was identified as the first date after wastewater onset for which only 50% of samples during a 14-d look back period had concentrations over or equal to 2,000 cp/g. 2000 cp/g was chosen as it represents approximately two times the detection limit, as described previously. ^21^ Wastewater event onset and offset dates were identified for the entire state and county as follows. The date of wastewater onset was determined as a date when 50% or more of the individual WWTPs in the state or country were in onset, as described above for the individual plants and offset as the date after onset for which less than 50% of the sites in the state or nation were in onset. For this analysis, an individual plant was not given an onset or offset designation for the first 14 days of data availability.

We compared epidemic peaks as defined by diagnostic and hospital data to the state-aggregated peaks in RSV wastewater concentrations. Peaks of clinical data were determined as the week with the highest values (last date of MMWR weak) whereas wastewater peaks were identified as the day with highest value of RSV_PMMoV_w_trimmed_s.

## Results

Over the study period of 1 January 2022 to 31 July 2023, wastewater data was available from 176 WWTPs in 34 states (Table 1). Population coverage of the wastewater catchment areas varied from 0.5% to 59.5% with a median (interquartile range, IQR) state-aggregated WWTP population of 324045 (119000, 693000).

**Table 1.** Spearman’s rho and p values for correlations between state-aggregated wastewater concentrations and positivity rate and hospitalization rate. Hospitalization rate is only available for a small subset of states with wastewater data and a blank cell indicates there was no hospitalization data available for that state. NA indicates that a rho could not be calculated due to lack of variation in one of the two variables. N is the number of data points used in the correlations.

| State | Rho between WW and positivity rate | P value of rho between WW and positivity rate | Rho between WW and hospitalization rate | P value of rho between WW and hospitalization rate | N |
| --- | --- | --- | --- | --- | --- |
| Alabama | 0.75 | $4.42 \times 10^{-10}$ | | | 49 |
| Alaska | 0.51 | 0.164 |  |  | 9 |
| Arkansas | NA | NA |  |  | 13 |
| California | 0.89 | $3.04 \times 10^{-29}$ | 0.90 | $1.98 \times 10^{-16}$ | 81 |
| Colorado | 0.8 | $3.54 \times 10^{-15}$ | 0.80 | $6.8 \times 10^{-8}$ | 62 |
| Delaware | NA | NA |  |  | 24 |
| Florida | 0.81 | $1.18 \times 10^{-16}$ | | | 68 |
| Georgia | 0.82 | $1.29 \times 10^{-14}$ | 0.86 | $3.8 \times 10^{-9}$ | 56 |
| Hawaii | 0.09 | 0.783 |  |  | 11 |
| Idaho | 0.75 | $1.51 \times 10^{-14}$ | | | 73 |
| Illinois | 0.8 | $1.27 \times 10^{-12}$ | | | 51 |
| Indiana | 0.71 | $1.03 \times 10^{-8}$ | | | 49 |
| Iowa | 0.93 | $3.00 \times 10^{-14}$ | | | 32 |
| Kansas | 0.81 | $4.51 \times 10^{-13}$ | | | 51 |
| Kentucky | 0.46 | $3.52 \times 10^{-05}$ | | | 73 |
| Maine | 0.8 | $3.43 \times 10^{-11}$ | | | 46 |
| Maryland | 0.83 | $3.02 \times 10^{-9}$ | 0.93 | $1.94 \times 10^{-7}$ | 32 |
| Massachusetts | 0.82 | $6.47 \times 10^{-9}$ | | | 32 |
| Michigan | 0.71 | $2.31 \times 10^{-11}$ | 0.89 | $2.1 \times 10^{-12}$ | 66 |
| Minnesota | 0.26 | 0.0757 | 0.31 | 0.14 | 47 |
| Nevada | 0.36 | 0.154 |  |  | 17 |
| New Hampshire | 0.77 | $3.96 \times 10^{-9}$ | | | 41 |
| New Jersey | 0.74 | $8.59 \times 10^{-10}$ | | | 50 |
| North Carolina | 0.86 | $7.92 \times 10^{-15}$ | | | 48 |
| Ohio | 0.8 | $4.92 \times 10^{-8}$ | | | 32 |
| Pennsylvania | 0.8 | $1.74 \times 10^{-12}$ | | | 51 |
| South Dakota | NA | NA |  |  | 13 |
| Tennessee | NA | NA | -0.5 | 0.66 | 6 |
| Texas | 0.42 | 0.000259 |  |  | 73 |
| Utah | 0.7 | $1.51 \times 10^{-7}$ | 0.93 | $2.7 \times 10^{-9}$ | 44 |
| Vermont | 0.56 | 0.00968 |  |  | 20 |
| Virginia | 0.3 | 0.0969 |  |  | 32 |
| West Virginia | NA | NA |  |  | 15 |
| Wisconsin | NA | NA |  |  | 5 |

### QA/QC

All wastewater measurements positive and negative controls were positive and negative respectively indicating acceptable assay performance and no contamination across the entire study. Median (IQR) BCoV recoveries across all samples were 1.1 (0.83, 1.2) indicating good recovery across the samples. Recoveries higher than 1 are likely a result of uncertainties in the measurement of BCoV added to the buffer matrix. Additional details of reporting outlined in the Environmental Microbiology Minimal Information (EMMI) reporting guidelines are provided elsewhere.^27^

### RSV measurements and correlations with clinical RSV data

RSV measurements at individual plants varied from non-detect to 9.4×10^6^ cp/g dry weight (median across all measurements was 1.1×10^3^ cp/g). Time series plots of RSV measurements from all WWTPs are provided in the supporting material (Figure S2). We generally found undetectable levels in the late spring and summer, with peak values in the winter in line with general expectations for seasonality of RSV in the Northern hemisphere.^10^

Wastewater concentrations paralleled clinical surveillance data. State-aggregated population-weighted average RSV/PMMoV (RSV_PMMoV_w_trimmed_s) was calculated for each state for which wastewater data were available (N=34), and Spearman’s rho was determined between the RSV_PMMoV_w_trimmed_s and state-aggregated positivity rate. Rho for six states could not be calculated because available wastewater values were constant; and rho was not statistically significant for five states (Table 1). The vast majority of the states for which rho was not statistically significant or could not be calculated had RSV wastewater data available only after late spring / early summer of 2023, after the RSV season had passed (Figures S2 and S3, Table S1). Rhos for the remaining states were statistically significant (rho between 0.42 and 0.93, p<0.001). Both wastewater and hospitalization rate data were available for 8 states and so rho was calculated to assess their associations (Table 1). Rho varied between 0.80 and 0.93 (p<0.001) for 6 states. A rho could not be calculated for one of the remaining states (all available wastewater values during the data collection period were non-detect), and the rho was insignificant for the other state (wastewater data collection started in June 2023, after RSV season). Nationally aggregated RSV/PMMoV (RSV_PMMoV_w_trimmed_n) was positively associated with nationally aggregated positivity and hospitalization rates (rho = 0.73, p = 2.2×10^−16^ and rho = 0.84, p=1.35×10^−10^, respectively).

### Onset, offset and peak of RSV season

Wastewater data was available before clinical onset of RSV season in 14 states (CA, CO, FL, ID, IL, IN, KS, KY, MI, ME, NH, NJ, UT, TX, Figure S2). In these states, clinical onset of RSV season occurred between 30 April 2022 and 12 November 2022, and clinical offset occurred between 24 December 2022 and 4 March 2023, depending on state (Table 2). Duration of RSV epidemic ranged from 14 to 39 weeks across states; longer duration was generally observed in states with earlier onset. Most (12 of 14) states had one RSV onset during this time period; UT and KY had multiple clinical onset and offset events (Figure 2, Table 2). For KY, the start of the first onset event was used as the date of clinical onset as the positivity rate tended to have an increasing slope with values not equal to zero between onset events. UT had onset events punctuated by multiple weeks of a zero value, which we suspect may be caused by a low number of tests administered.^32^ Therefore, for UT, we considered the clinical onset date to be that which occurred prior to the peak in positivity rates.

**Figure 1.**
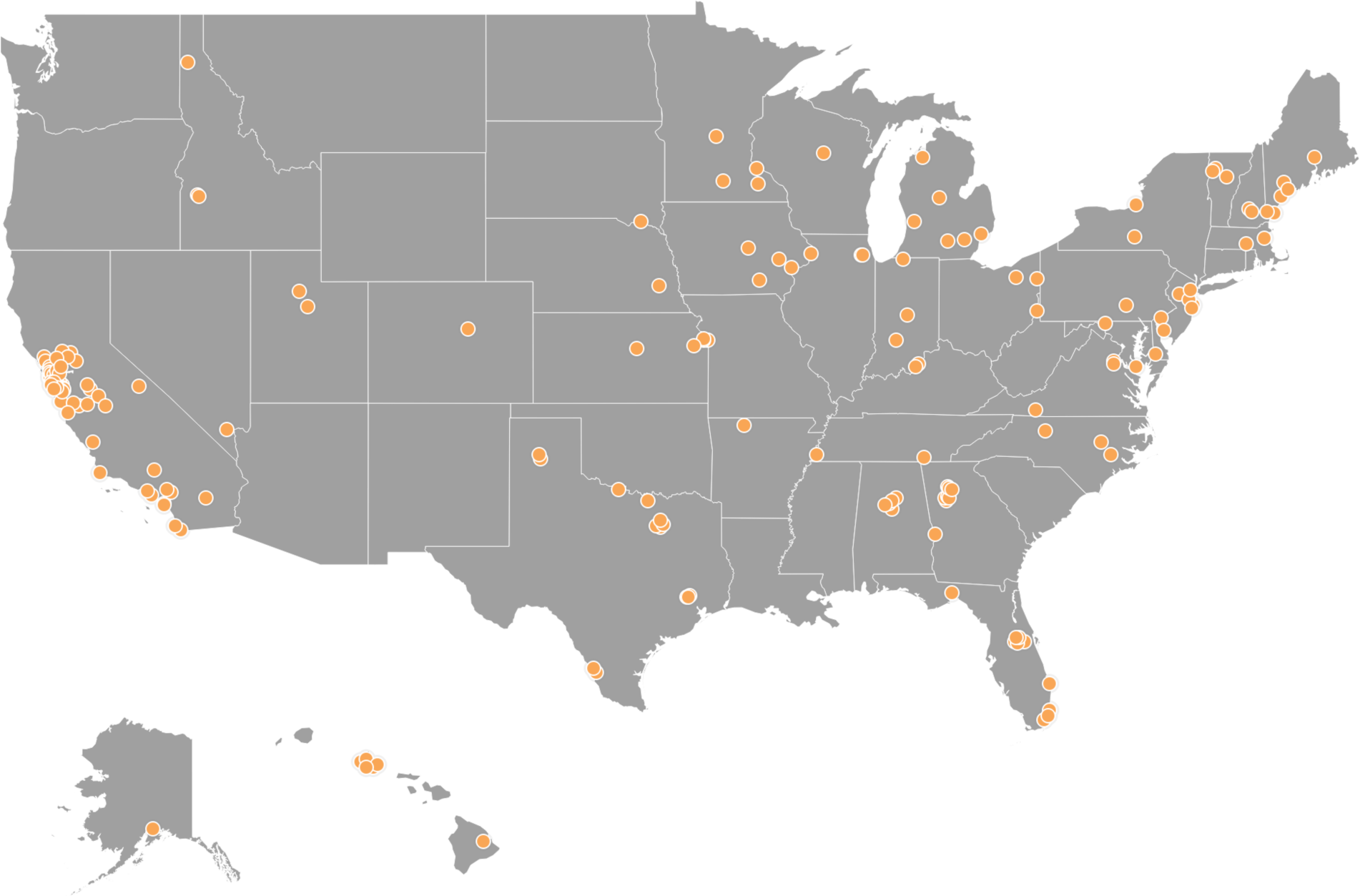
Location of WWTPs with RSV RNA data included in the paper. Each orange dot represents the location of each WWTP.

**FIGURE 2.**
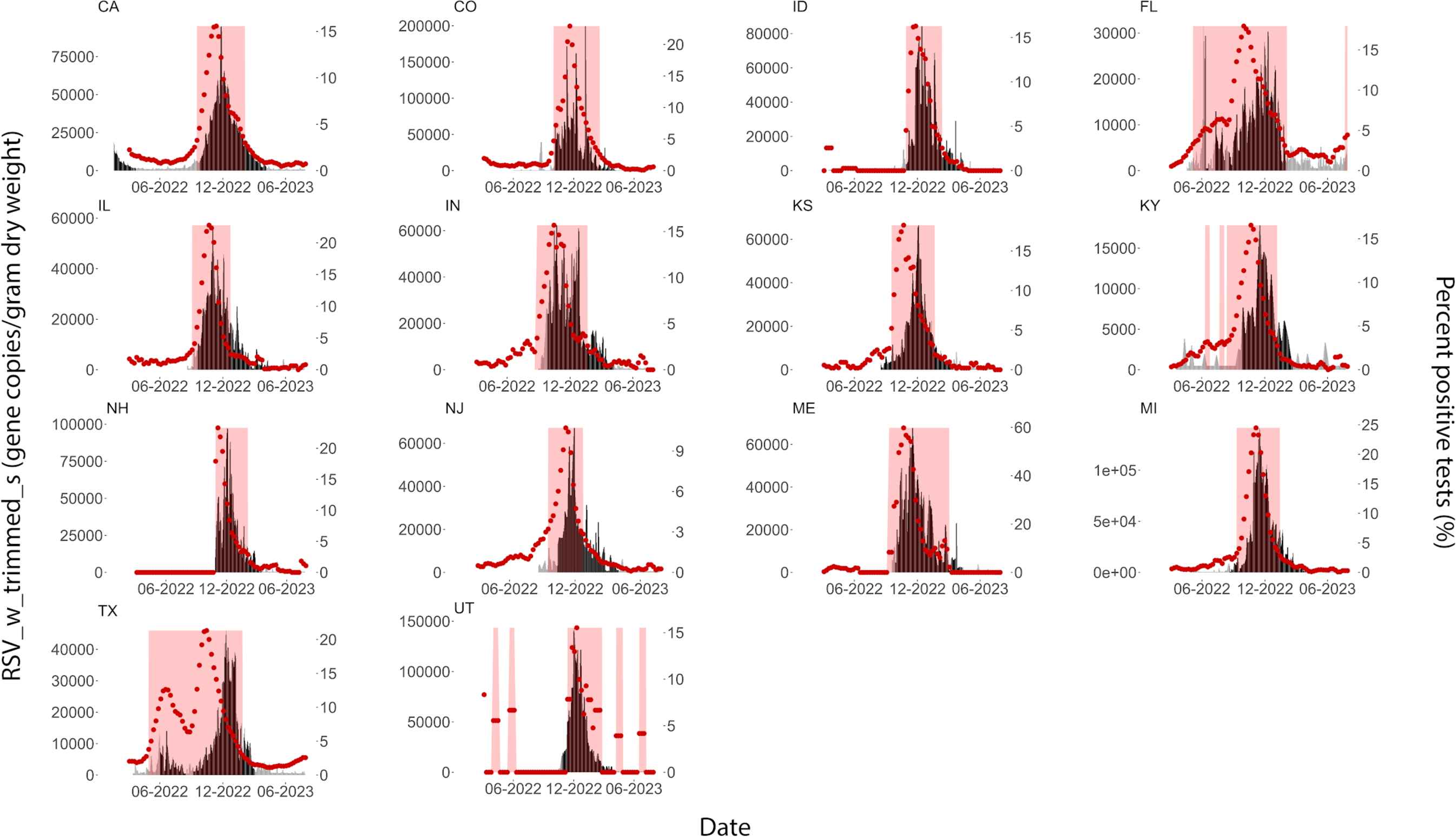
State-aggregated concentrations of RSV RNA (RSV_w_trimmed_s, shaded area, left axis) and positivity rates (red dots, right axis). Red background shading indicates periods between clinical onset and offset. Black fill below the wastewater concentrations indicates periods of wastewater event onset, whereas grey fill indicates time periods when wastewater events are not in onset.

**Table 2.** For each of 14 states with both wastewater and clinical data for the duration of the 2022-2023 RSV season, we show the dates of clinical onset and offset; if there are more than one, they are all given separated by a semicolon and a bolded date is the one used for Figure 3 and in the text to describe the state of the RSV season. The weekly median RSV_PMMoV_w_trimmed_s during the MMWR week of clinical onset and offset is provided; the value is unitless. “WW onset,offset” provides the dates of wastewater event onset and offset; if more than one occurred, then the dates are separated by semicolons. The date of onset of the first wastewater event is used in Figure 3 and in the text. Date of the peak value for hospitalization (hosp.) rate, positivity rate, and wastewater (RSV_PMMoV_w_trimmed_s) are provided. All dates are month/day/year format.

| State | Clinical onset,offset | WW median at onset (x 10^6) | WW median at offset (x 10^6) | WW onset,offset | Date of peak hosp. rate | Date of peak positivity rate | Date of peak WW |
| --- | --- | --- | --- | --- | --- | --- | --- |
| California | 9/17/2022, 2/4/2023 | 5.5 | 12.8 | 9/25/22, 4/6/23 | 12/3/2022 | 11/12/2022 | 11/27/2022 |
| Colorado | 10/1/2022, 2/18/2023 | 10.5 | 17.5 | 10/4/22, 4/9/23 | 11/12/2022 | 11/19/2022 | 11/09/2022 |
| Florida | 5/7/2022, 2/4/2023 | 8.7 | 8.6 | 6/21/22, 8/9/22; 8/19/22, 1/31/23 |  | 10/1/2022 | 6/11/2022 |
| Idaho | 10/29/2022, 2/11/2023 | 8.3 | 47.3 | 10/29/22, 4/11/23 |  | 11/26/2022 | 12/14/2022 |
| Illinois | 9/3/2022, 12/24/2022 | 2.2 | 1.9 | 9/20/22, 4/25/23 |  | 10/22/2022 | 10/30/2022 |
| Indiana | 8/27/2022, 1/21/2023 | 8.0 | 3.0 | 9/27/22, 4/7/23 |  | 10/15/2022 | 11/8/2022 |
| Kansas | 9/17/2022, 1/21/2023 | 7.2 | 31.6 | 8/17/22, 3/15/23 |  | 10/22/2022 | 12/2/2022 |
| Kentucky | **6/11/2022**, 6/25/2022; 7/23/2022, 7/30/2022; 8/06/2022, **1/7/2023** | 1.8 | 16.5 | 9/26/22, 2/21/23 |  | 10/22/2022 | 11/15/2022 |
| Maine | 9/17/2022, 3/4/2023 | 3.0 | 12.2 | 9/28/22, 4/10/23 |  | 10/22/2022 | 11/12/2022 |
| Michigan | 9/10/2022, 1/14/2023 | 2.0 | 92.2 | 8/20/22, 3/12/23 | 11/12/2022 | 11/5/2022 | 11/17/2022 |
| New Hampshire | 10/29/2022, 2/4/2023 | 85 | 58.6 | 10/28/22, 3/17/23 |  | 11/5/2022 | 12/13/2022 |
| New Jersey | 9/17/2022, 12/24/2022 | 4.6 | 71.4 | 10/13/22, 4/2/23 |  | 11/5/2022 | 12/17/2022 |
| Texas | 4/30/2022, 1/28/2023 | 1.6 | 13.6 | 6/2/22, 8/16/22; 9/7/22, 3/5/23 |  | 10/15/2022 | 12/10/2022 |
| Utah | 4/02/2022, 4/23/2022; 5/21/2022, | 27 | 7.9 | 10/26/22, 4/5/23 | 11/26/2022 | 12/10/2022 | 12/4/2022 |
|  | 6/11/2022;<br>**11/12/2022,**<br>**2/25/2023** |  |  |  |  |  |  |

State weekly median PMMoV-normalized wastewater concentrations (RSV_PMMoV_w_trimmed_s) during the week of clinical onset ranged from 2.17×10^−6^ to 8.50×10^−5^ while the weekly median concentrations during the week of clinical offset ranged from 1.88×10^−6^ to 9.22×10^−5^, depending on state (Table 2). Weekly median non-normalized wastewater concentrations (RSV_w_trimmed_s) during the week of clinical onset ranged from 170 to 24,000 cp/g while the weekly median concentrations during the week of clinical offset franged from 2,400 to 39,000 cp/g (Table 2).

For the 14 states, the date of wastewater onset varied between 21 June to 29 October 2022, and date of offset varied between 31 January to 25 April 2023 (Table 2), with the duration of the event between 20 and 37 weeks. Most (12 of 14) of the state wastewater data showed one RSV wastewater event over the study period (Figure 2, Table 2). Two states (FL and TX) had multiple RSV wastewater events. TX had two, one between early June and mid August 2022, and the other between early September 2022 and early May 2023. FL had three RSV wastewater events: one between late-June and mid-July 2022, one between late July and early August 2022, and one between mid August and late January 2023. The date of wastewater onset for TX and FL was the onset date of the first wastewater event.

Peaks in state-aggregated population weighted average RSV/PMMoV (RSV_PMMoV_w_trimmed_s) across the 14 states ranged from 30 October to 17 December 2022, with FL peak being in mid-June (Table 2). Peaks in hospitalization rates, for the 4 of 14 states with available hospitalization rate data, were within the same week as the peak in wastewater data (Table 2). Peaks in positivity rate data were generally earlier than the peaks in the wastewater variable. For three states (FL, CO, and UT, wastewater peaked approximately 1-16 weeks earlier than positivity rate data. In the remaining 11 states, positivity rates peaked before wastewater (median 3 weeks earlier, range1-8 weeks earlier).

The 2022-2023 RSV season demonstrated a consistent geo-temporal pattern of wastewater onset (Figure 3). Wastewater onset occurred earliest in the south in Texas and Florida. Then, wastewater onset moved north and west almost uniformly, with the midwest (Kansas and Michigan) first followed closely by the remaining states where wastewater onset occurred nearly in coherence. Clinical onset followed a similar pattern with clinical onset occurring in Texas and Florida first, followed by a move of onset both west and north.

**FIGURE 3.**
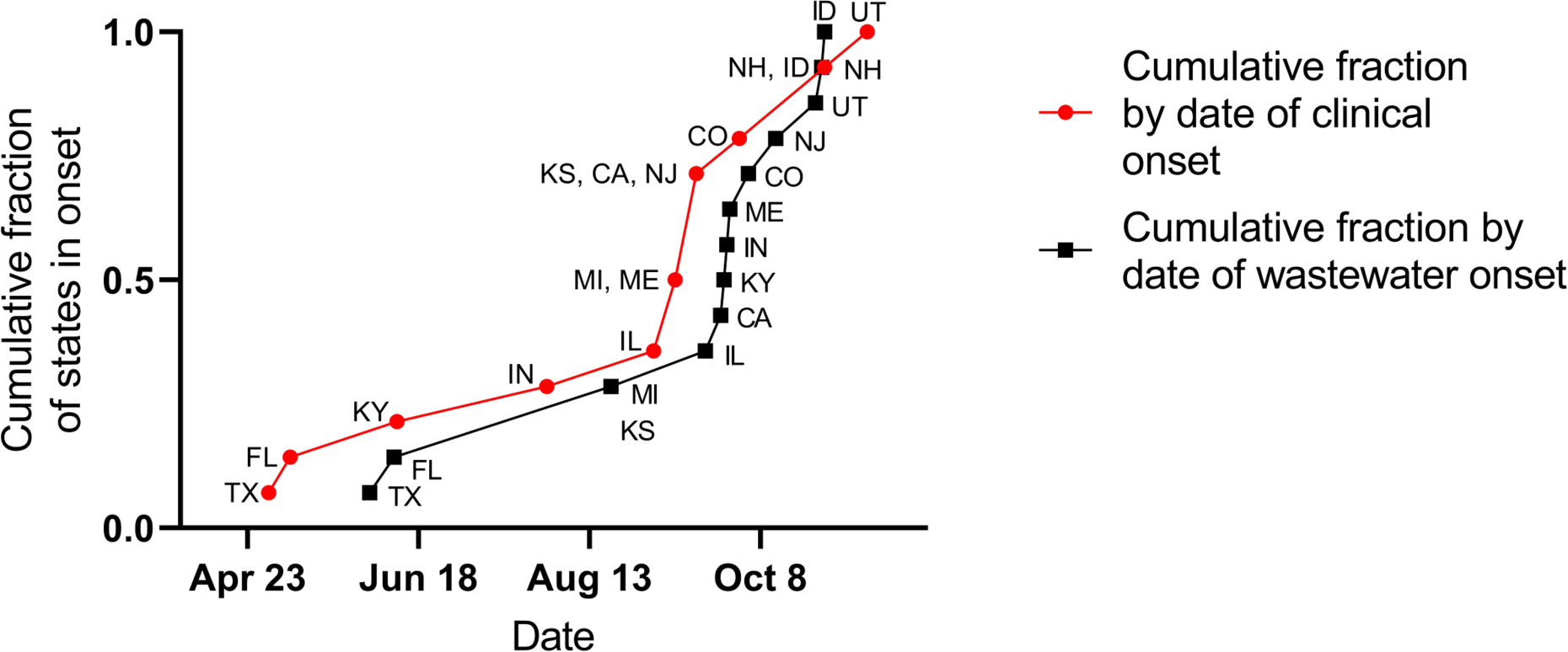
Cumulative fraction of 14 states (y-axis) in RSV onset, as determined by clinical positivity rates in red and by wastewater concentrations in black. The date of clinical onset (red) or wastewater event onset (black) is shown on the x-axis. Each symbol is labeled with the state.

## Discussion

We show that RSV RNA concentrations in wastewater collected throughout the United States correlate to both positivity rates and hospitalizations nationally and at the state level, as shown for single or small groups of wastewater treatment plants in previous studies.^15–17,19–21^ Sites varied from urban to rural areas, spanned climates from tropical to continental,^33^ and represented populations served from 3000 to 4 million per site with overall coverage of 0.05% to 59.5% of the state population. Despite significant diversity in site characteristics and coverage from state to state, correlations to clinical positivity rates and hospitalizations were positive and significant. Hospitalization peaks largely matched wastewater peaks temporally, suggesting the value of wastewater monitoring for informing hospital staffing and public health notifications. For the four states with sufficient hospitalization and wastewater data, the peaks of both wastewater and hospitalizations all fell within an 8 day range of each other.

We determined the onset of RSV season using two independently measured signals, clinical positivity rates and wastewater measurements. It is important to note that clinical onset date is identified retroactively, determined after 2 consecutive weeks of clinical positivity rates greater than 3%.^24,34^ By comparison, the wastewater event onset algorithm determines the onset of RSV wastewater events in real time. For 3 states, wastewater event onset occurred the same week as clinical onset; for 3 states, wastewater event onset occurred 2 to 4 weeks before clinical onset; for 7 states, wastewater event onset commenced 1 to 15 weeks after clinical onset. Kentucky (KY) was the state for which the wastwater event onset occurred 15 weeks after clinical onset; in this case, clinical onset was identified in June 2022 while wastewater onset was identified in September 2022. RSV dynamics in the 2022-2023 season were likely affected by changes in susceptibility of individuals and early start to RSV activity in some locations likely due to the suppression of RSV activity during the first two years of COVID-19 pandemic.

While wastewater data were well correlated with positivity rate data, peaks and onset determined using positivity rate data and wastewater data were not always aligned. There are several explanations for this. Wastewater concentrations are controlled by inputs to the system from every individual shedding RSV RNA, including those with severe or mild symptoms and those who are both symptomatic and asymptomatic. There are very limited data on longitudinal RSV shedding in human excretions, and it is possible prolonged shedding in convalescing or recovered individuals occurs.^35^ Lowry et al. found not a single study is available that quantifies concentrations of RSV RNA in excretions that contribute to wastewater.^35^ Positivity rate measures occurrence of RSV infections in individuals who are ill enough to seek medical care, most commonly at an early and acute stage of infection and may include infants in diapers who are less likely to contribute to wastewater. While wastewater likely captures repeated inputs from infected individuals, diagnostic laboratory tests are generally performed and entered into surveillance systems once. Therefore, the differences in onset and peaks in the two measures could reflect different patterns of incident symptomatic, severe RSV (reflected by the positivity rate data) and epidemic trajectory of total viral burden (reflected by the wastewater data) as well as differences in the contributing populations.

An alternate explanation for the lack of agreement in peaks and onset according to wastewater and positivity rate data streams are limitations and biases associated with each. The majority of RSV infections are unrecognized and are not captured by clinical surveillance systems. In a prospective surveillance study of more than 5000 children in the U.S. with an acute respiratory infection, only 45% of inpatient RSV infections– and a mere 3% of outpatient infections– received the diagnosis RSV-associated illness.^6^ Publicly available state-aggregated positivity rate data can also be affected by the number of tests administered. In our data series, some states reported as few as 1 performed test per week, which limits the ability to determine a reliable estimate of positivity rate. In addition, clinical testing rates are not temporally uniform and tend to fluctuate according to the clinical landscape and clinicians’ awareness of ongoing RSV circulation. For example, on the date of clinical onset in IL, 321 weekly tests were performed, while on the date of offset, 1370 weekly tests were performed. Additionally, publicly available positivity rate data was only available weekly, as 3-week averages limit the resolution at which changes in positivity rates can be identified. Positivity rate data are generated by clinical laboratories participating in NREVSS which is a passive, voluntary program. The number of laboratories participating in NREVSS varies by state (N=1 to 29 per state), and the exact laboratories contributing positivity rate data at any time are not publicly available and may change over time.^34^ Similarly, the number of WWTP participating in wastewater surveillance during the study period differed across states (N=1 to 57 per state). The spatial coverage of the participating laboratories testing for RSV for NREVSS likely does not exactly match the spatial coverage of the WWTP in the state (Figure S4).

We posit that wastewater measurements of RSV can augment traditional RSV season indicators like hospitalization and positivity rates, and may provide complementary information about population-level RSV infections. Wastewater data are available in real time (as soon as 24 hours after sample collection) and are not subjected to delays associated with clinical data reporting.^15,36,37^. In addition, wastewater measurements are not subject to any clinical testing biases typically present due to socioeconomic factors or testing availability.^37,38^ Importantly, wastewater data can be available at locations where or times when traditional disease metrics like positivity and hospitalization rates, are not available offering the ability to provide public health resources at appropriate spatial and temporal scales, and providing insights into disease epidemiology uniquely available through wastewater.

The PMMoV-normalized RSV RNA concentration in wastewater during clinical onset and offset was of a similar magnitude across states and the country suggesting a simple threshold may be useful for identifying dates of clinical onset and offset, using wastewater. The onset and offset algorithms used in this study represent an example approach for identifying significant emergence of RSV RNA in wastewater, but different wastewater event onset and offset thresholds or algorithms that are fit for their specific public health or other stakeholder use-cases can be developed.

More work is needed to better understand shedding of RSV in human excretions to provide a linkage between wastewater concentrations and disease occurrence in the contributing population. Until such data are available, the data provided herein suggests a strong relationship between RSV RNA in wastewater solids across the United States, and more traditional RSV indicators. Future work can use wastewater to explore the relative importance of RSV A and RSV B.^39^ While most clinical tests do not distinguish between the two, some evidence suggests there may be differences in disease severity for the different subtypes.^40^

## Conclusion

Wastewater concentrations of RSV RNA can complement traditional clinical indicators of disease and inform public health and clinical decision making. Wastewater may be useful in the near future as an unbiased metric to assess the effectiveness of new RSV vaccines and other immunization strategies. Three new immunization strategies are now available against RSV – monoclonal antibody for children under 2, maternal vaccination with bivalent RSV prefusion F protein-based (RSVPreF) vaccine, and active vaccination of individuals 60 and over with RSVPreF or adjuvant RSVPreF3. The protection afforded by these methods wanes with time, which makes administration relative to RSV season onset crucial. Wastewater data presents a unique opportunity to monitor when vaccine campaigns should be promoted, the subsequent effectiveness of immunization efforts through careful comparison of immunization levels and wastewater concentrations, and possible antigenic changes in circulating virus as population immunity increases across the US.

## Data Availability

All data produced are available online at https://purl.stanford.edu/hd388xb1982

## Acknowledgements

We acknowledge the numerous people who contributed to wastewater sample collection. Graphical abstract created with BioRender.com.

## Notes

### Competing Interest Statement

The authors have declared no competing interest.

### Funding Statement

This study was funded by the CDC Foundation and the Sergey Brin Family Foundation

### Author Declarations

The study used ONLY openly available human data that were originally located at https://www.cdc.gov/rsv/research/rsv-net/dashboard.html and https://www.cdc.gov/surveillance/nrevss/rsv/index.html

