## Supporting Information for "Observations of respiratory syncytial virus (RSV) nucleic-acids in wastewater solids across the United States in the 2022-2023 season: Relationships with RSV infection positivity and hospitalization rates"

Number of pages: 47
Number of Tables: 2
Number of Figures: 3

### Supplementary Methods

**Additional details on pre-analytical methods.** The pre-analytical methods have been registered at protocols.io[1] and described in other publications[2]. Pre-analytical processing occurred as soon as the samples were received at the laboratory. In brief, wastewater solids are dewatered using centrifugation. Thereafter, one aliquot is taken to measure the dry weight using an oven. Another aliquot is added to bovine-coronavirus-spiked DNA/RNA shield (Zymo, Irvine CA) so the final concentration is ~75 mg/ml; at this concentration, we observe minimal inhibition of the assays.[3] The solution is then homogenized using grinding balls, and then centrifuged again. The nucleic-acids are then extracted from an aliquot of the supernatant using a commercial extraction kit. This is done in 6 or 10 replicates so that there are 6 or 10 distinct nucleic-acid extractions from each solids sample. The only sites that used 10 replicates are Gilroy, CA; Oceanside San Francisco, CA; Palo Alto, CA; Redwood City, CA; Sacramento, CA; San Jose, CA; Southeast San Francisco, CA; and Sunnyvale, CA. The rest of the WWTPs used 6 replicates.

**Additional details on the analytical methods.** Nucleic-acids were processed immediately with no storage. We measured concentrations of the RSV N gene using a primer and probe set that has been previously published and applied to wastewater[4] using droplet digital RT-PCR. The nucleic-acid extracts were used as template neat (not diluted). Each nucleic-acid extract was run in its own well so that 6 or 10 replicate wells were run per sample. The RSV assay was multiplexed with other assays, and those assays changed over the period of the project as public health needs changed and new research to support the monitoring of additional disease targets became available; the assays RSV was multiplexed with are provided in Figure S1. PMMoV and BCoV were assayed using the same methods as previously described[3]; nucleic-acids were diluted 1:100 as template; two or 10 wells were run per sample; the sites where 10 wells were run are the same as those mentioned in the previous section. The specific methods have been described in detail elsewhere[3]. The replicate wells were merged for analysis and the methods for thresholding were described by Boehm et al.[3]

**QA/QC.** Extraction positive and negative controls, as well as RT-PCR positive and negative controls were included on each plate. All positive and negative controls were positive and negative, respectively. More information on the QA/QC are provided by Boehm et al.[3] regarding the preparation of the controls. In order for a sample to be counted as positive, it had to have 3 positive droplets across the merged wells. This is equivalent to a concentration of about 500-1000 copies per gram dry weight; the range reflects the varied dry weights of the solids, and whether 6 or 10 wells were used for the sample.

**Table S1.** Details of the 176 WWTPs included in this study.

The state where the WWTP is located, additional details regarding HHS region, plant location, site name, sample type, reported population served, and sample RSV descriptive statistics. ND: Non-detect, Min: Minimum, Max: Maximum, Med: Median. If the sample type is listed as "Solids" then that means the site provided samples of settled solids from the primary clarifier. If the sample type is labeled as "Solids (Settling w/ Imhoff cones on site)" the solids were obtained from raw influent using an Imhoff cone on site. If the sample type is labeled "Liquid", then the solids were obtained in the laboratory by allowing the raw influent to settle for 10-15 mins, and using a serological pipette to aspirate the settled solids into a falcon tube. The dates are in month/day/year format.

| State | Plant | Site Name | Sample Type | Population Served | Sample Count | Sample Start | Sample End | Min RSV (cp/g) | Max RSV (cp/g) | Med RSV (cp/g) |
|---|---|---|---|---|---|---|---|---|---|---|
| Alabama | Bessemer, AL | Valley Creek Water Reclamation Facility | Liquids | 225,000 | 148 | 8/17/22 | 7/26/23 | ND | 108361 | 4504 |
| | Cahaba River, Birmingham, AL | Cahaba River Water Reclamation Facility | Liquids | 95,000 | 127 | 8/17/22 | 7/26/23 | ND | 265111 | 5773 |

| | | | | | | | | | | |
|---|---|---|---|---|---|---|---|---|---|---|
| | Fultondale, AL | Five Mile Creek Water Reclamation Facility | Liquids | 77,000 | 113 | 8/24/22 | 7/26/23 | ND | 37431 | ND |
| | Pinson, AL | Turkey Creek Water Reclamation Facility | Liquids | 30,000 | 115 | 8/15/22 | 7/26/23 | ND | 64275 | ND |
| | Village Creek, Birmingham, AL | Village Creek Water Reclamation Facility | Liquids | 200,000 | 142 | 8/18/22 | 7/20/23 | ND | 123112 | ND |
| Alaska | Anchorage, AK | John M. Asplund Water Pollution Control Facility | Liquids | 220,000 | 22 | 5/30/23 | 7/18/23 | ND | 26135 | ND |
| Arkansas | Harrison, AR | City of Harrison Wastewater Treatment Plant | Liquids | 15,000 | 39 | 4/26/23 | 7/28/23 | ND | ND | ND |

| | | | | | | | | | | |
|---|---|---|---|---|---|---|---|---|---|---|
| California | Coastal, Laguna Niguel, CA | Coastal Treatment Plant | Liquids | 48,000 | 93 | 12/23/22 | 7/26/23 | ND | 91888 | 500 |
| | Contra Costa County, CA | Central Contra Costa Sanitary District | Solids | 484,800 | 189 | 3/24/22 | 7/24/23 | ND | 161227 | 3235 |
| | Davis, CA | City of Davis Wastewate r Treatment Plant | Solids | 68,000 | 446 | 1/9/22 | 7/26/23 | ND | 114816 | 1684 |
| | Esparto, CA | Esparto Wastewate r Treatment Facility | Liquids | 4,006 | 93 | 12/5/22 | 7/26/23 | ND | 112029 | ND |
| | Fairfield, CA | Fairfield-Su isun Sewer District | Solids | 155,000 | 128 | 9/30/22 | 7/26/23 | ND | 83389 | 4430 |

| | | | | | | | | | | |
|---|---|---|---|---|---|---|---|---|---|---|
| | Fremont, CA | [Fremont Basin] - Raymond A. Boege Alvarado WWTP | Liquids | 229,476 | 69 | 11/3/22 | 7/25/23 | ND | 82827 | 6757 |
| | Gilroy, CA | South County Regional Wastewater Authority | Solids (Settling w/ Imhoff cones on site) | 110,338 | 565 | 1/9/22 | 7/28/23 | ND | 250039 | 2344 |
| | Half Moon Bay, CA | Sewer Authority Mid-Coastside | Liquids | 28,000 | 172 | 4/29/22 | 7/26/23 | ND | 201535 | ND |
| | Hollister, CA | City of Hollister Domestic Water Recycling Facility | Liquids | 42,000 | 123 | 9/16/22 | 7/26/23 | ND | 647946 | 3086 |
| | Indio, CA | Valley Sanitary District | Solids | 91,765 | 144 | 8/26/22 | 7/26/23 | ND | 114431 | 2325 |

| | | | | | | | | | | |
|---|---|---|---|---|---|---|---|---|---|---|
| | JB Latham, Laguna Niguel, CA | JB Latham Treatment Plant | Liquids | 120,000 | 93 | 12/23/22 | 7/26/23 | ND | 89784 | ND |
| | Lancaster, CA | Lancaster Water Reclamation Plant | Liquids | 200,000 | 134 | 9/15/22 | 7/25/23 | ND | 208991 | 3693 |
| | Las Gallinas, San Rafael, CA | Las Gallinas Valley Sanitary District | Liquids | 30,000 | 151 | 8/10/22 | 7/24/23 | ND | 326267 | 6680 |
| | Lompoc, CA | Lompoc Regional Wastewater Reclamation Plant | Liquids | 69,290 | 152 | 8/3/22 | 7/21/23 | ND | 225788 | ND |
| | Los Angeles County, CA | Joint Water Pollution Control Plant | Liquids | 3,500,000 | 217 | 3/2/22 | 7/26/23 | ND | 75718 | ND |
| | Los Angeles, CA | Hyperion Water Reclamation Plant (HWRP) | Liquids | 4,000,000 | 142 | 8/30/22 | 7/26/23 | ND | 125026 | 7246 |

| | | | | | | | | | | |
|---|---|---|---|---|---|---|---|---|---|---|
| | Los Banos, CA | Los Banos Wastewater Treatment Plant | Liquids | 42,000 | 102 | 12/6/22 | 7/25/23 | ND | 186224 | ND |
| | Madera, CA | City of Madera, Wastewater Treatment Plant | Solids | 67,944 | 58 | 3/8/23 | 7/26/23 | ND | 8244 | ND |
| | Marina, CA | Monterey One Water - Regional Treatment Plant | Liquids | 262,000 | 98 | 11/29/22 | 7/27/23 | ND | 154473 | 2879 |
| | Mammoth, CA | Mammoth Community Water District | Liquids | 35,000 | 43 | 3/17/23 | 6/28/23 | ND | 25284 | ND |
| | Merced, CA | Merced Wastewater Treatment Plant | Solids (Settling w/ Imhoff cones on site) | 91,000 | 214 | 1/8/22 | 7/26/23 | ND | 92791 | 2083 |

| | | | | | | | | | | |
|---|---|---|---|---|---|---|---|---|---|---|
| | Mill Valley, CA | Sewerage Agency of Southern Marin Wastewater Treatment Plant | Solids | 30,000 | 96 | 12/15/22 | 7/25/23 | ND | 72795 | ND |
| | Modesto, CA | Modesto's Sutter Primary Treatment Facility | Solids | 230,000 | 209 | 1/7/22 | 7/24/23 | ND | 163909 | ND |
| | Napa, CA | Soscol Water Recycling Facility | Solids | 83,300 | 125 | 9/28/22 | 7/24/23 | ND | 127519 | 16535 |
| | Newark, CA | [Newark Basin] - Raymond A. Boege Alvarado WWTP | Liquids | 47,229 | 70 | 11/8/22 | 7/25/23 | ND | 116431 | ND |
| | Novato, CA | Novato Sanitary District | Liquids | 53,000 | 166 | 6/22/22 | 7/26/23 | ND | 208474 | 4191 |

| | | | | | | | | | | |
|---|---|---|---|---|---|---|---|---|---|---|
| | Oakland, CA | East Bay Municipal Utility District | Solids | 740,000 | 219 | 3/3/22 | 7/24/23 | ND | 64747 | ND |
| | Oceanside, San Francisco, CA | Oceanside Water Pollution Control Plant | Solids | 250,000 | 540 | 1/9/22 | 7/27/23 | ND | 128349 | 3111 |
| | Ontario, CA | Regional Water Recycling Plant No.1 (RP-1) | Solids | 890,000 | 188 | 4/27/22 | 7/24/23 | ND | 213511 | ND |
| | Pacifica, CA | Calera Creek Water Recycling Plant | Liquids | 40,000 | 116 | 10/19/22 | 7/26/23 | ND | 310047 | 3366 |
| | Palo Alto, CA | Palo Alto Regional Water Quality Control Plant | Solids | 236,000 | 566 | 1/8/22 | 7/28/23 | ND | 162100 | 4641 |

| | | | | | | | | | | |
|---|---|---|---|---|---|---|---|---|---|---|
| | Paso Robles, CA | City of Paso Robles Wastewater Treatment Plant | Solids | 31,037 | 217 | 3/3/22 | 7/25/23 | ND | 162084 | 2217 |
| | Petaluma, CA | Ellis Creek Water Recycling Facility | Liquids | 65,000 | 138 | 6/30/22 | 5/24/23 | ND | 181055 | 10328 |
| | Redwood City, CA | Silicon Valley Clean Water | Solids | 199,000 | 566 | 1/9/22 | 7/28/23 | ND | 151294 | 3720 |
| | Regional, Laguna Niguel, CA | Regional Treatment Plant | Liquids | 129,000 | 92 | 12/23/22 | 7/26/23 | ND | 135432 | ND |
| | Riverside, CA | Riverside Water Quality Control Plant | Liquids | 350,000 | 78 | 1/27/23 | 7/26/23 | ND | 38977 | ND |

| | | | | | | | | | | |
|---|---|---|---|---|---|---|---|---|---|---|
| | Sacramento, CA | Sacramento Regional Wastewater Treatment Plant | Solids | 1,480,000 | 566 | 1/9/22 | 7/28/23 | ND | 163276 | 2120 |
| | San Diego, CA | E.W. Blom Point Loma Wastewater Treatment Plant | Liquids | 2,200,000 | 144 | 8/10/22 | 7/26/23 | ND | 141475 | 10392 |
| | San Jose, CA | San Jose-Santa Clara Regional Wastewater Facility | Solids | 1,500,000 | 567 | 1/8/22 | 7/28/23 | ND | 119499 | 4361 |
| | San Leandro, CA | City of San Leandro Water Pollution Control Plant | Liquids | 50,000 | 128 | 9/29/22 | 7/25/23 | ND | 271002 | 6496 |

| | | | | | | | | | | |
|---|---|---|---|---|---|---|---|---|---|---|
| | San Mateo, CA | City of San Mateo & Estero M.I.D. Water Quality Control Plant | Solids | 150,000 | 151 | 7/11/22 | 7/26/23 | ND | 115936 | 2684 |
| | San Rafael, CA | Central Marin Sanitation Agency | Liquids | 104,250 | 88 | 8/24/22 | 7/25/23 | ND | 142503 | 7499 |
| | Santa Cruz County, CA | City of Santa Cruz WTF - County Influent | Solids (Settling w/ Imhoff cones on site) | 160,000 | 201 | 4/5/22 | 7/27/23 | ND | 170990 | ND |
| | Santa Cruz, CA | City of Santa Cruz WTF - City Influent | Solids (Settling w/ Imhoff cones on site) | 160,000 | 200 | 4/5/22 | 7/27/23 | ND | 9423484 | ND |
| | Santa Rosa, CA | City of Santa Rosa, Laguna Treatment Plant | Solids (Settling w/ Imhoff cones on site) | 230,000 | 146 | 8/15/22 | 7/23/23 | ND | 113775 | 4199 |

| | | | | | | | | | | |
|---|---|---|---|---|---|---|---|---|---|---|
| | Sausalito, CA | Sausalito-Marin City Sanitary District | Liquids | 18,000 | 122 | 8/15/22 | 7/25/23 | ND | 220907 | 12258 |
| | South San Diego, CA | South Bay International Wastewater Treatment Plant | Solids | 1,600,000 | 33 | 12/19/22 | 3/8/23 | ND | 35467 | 15474 |
| | Southeast San Francisco, CA | Southeast San Francisco | Solids | 750,000 | 426 | 5/22/22 | 7/28/23 | ND | 182315 | 3607 |
| | Sunnyvale, CA | City of Sunnyvale Water Pollution Control Plant | Solids | 153,000 | 562 | 1/9/22 | 7/28/23 | ND | 151556 | 4172 |
| | Turlock, CA | Turlock Regional Water Quality Control Facility | Liquids | 86,000 | 101 | 12/5/22 | 7/26/23 | ND | 144759 | ND |

| | | | | | | | | | | |
|---|---|---|---|---|---|---|---|---|---|---|
| | Union City, CA | [Union City Basin] - Raymond A. Boege Alvarado WWTP | Liquids | 68,150 | 70 | 11/3/22 | 7/25/23 | ND | 224665 | 5190 |
| | Vallejo, CA | Vallejo Flood and Wastewater District Wastewater Treatment Plant | Liquids | 121,000 | 130 | 9/22/22 | 7/25/23 | ND | 269453 | 8597 |
| | West Contra Costa County, CA | West County Wastewater District | Liquids | 100,000 | 142 | 5/12/22 | 7/25/23 | ND | 1544895 | 2911 |
| | West Railroad, San Rafael, CA | Central Marin Sanitation Agency - West Railroad | Liquids | 25,000 | 90 | 8/24/22 | 7/25/23 | ND | 129905 | 7054 |

| | | | | | | | | | | |
|---|---|---|---|---|---|---|---|---|---|---|
| | Windsor, CA | Windsor Wastewater Treatment, Reclamation, and Disposal Facility | Liquids | 28,000 | 85 | 12/19/22 | 7/26/23 | ND | 178031 | ND |
| | Winters, CA | Winters - East Street Pump Station | Liquids | 7,286 | 92 | 12/5/22 | 7/26/23 | ND | 285824 | ND |
| | Woodland, CA | Woodland Water Pollution Control Facility | Liquids | 59,000 | 99 | 12/5/22 | 7/26/23 | ND | 559544 | ND |
| Colorado | North, Parker, CO | Parker Water and Sanitation District North Water Reclamation Facility | Liquids | 35,000 | 178 | 5/18/22 | 7/24/23 | ND | 445180 | ND |

| | | | | | | | | | | |
|---|---|---|---|---|---|---|---|---|---|---|
| | South, Parker, CO | Parker Water and Sanitation District South Water Reclamation Facility | Liquids | 25,000 | 179 | 5/18/22 | 7/24/23 | ND | 541129 | ND |
| Delaware | Seaford, DE | Seaford Wastewater Treatment Facility | Solids | 13,172 | 71 | 2/10/23 | 7/26/23 | ND | 11258 | ND |
| Florida | Altamonte Springs, FL | Altamonte Springs Regional Water Reclamation Facility | Liquids | 95,000 | 107 | 10/26/22 | 7/26/23 | ND | 86028 | ND |
| | Eastern, Orange County, FL | Eastern Water Reclamation Facility | Solids (Settling w/ Imhoff cones onsite) | 195,299 | 185 | 4/5/22 | 7/25/23 | ND | 112437 | ND |

| | | | | | | | | | | |
|---|---|---|---|---|---|---|---|---|---|---|
| | Jupiter, FL | Loxahatchee River Environmental Control District | Liquids | 90,000 | 132 | 9/16/22 | 7/26/23 | ND | 51684 | ND |
| | Key Biscayne, FL | MDWASD Central District WWTP | Liquids | 829,725 | 67 | 1/24/23 | 7/11/23 | ND | 53099 | ND |
| | North Miami, FL | MDWASD North District WWTF | Liquids | 776,150 | 81 | 1/18/23 | 7/26/23 | ND | 11764 | ND |
| | Northwest, Orange County, FL | Northwest Water Reclamation Facility | Solids (Settling w/ Imhoff cones onsite) | 66,690 | 187 | 4/5/22 | 7/27/23 | ND | 119608 | ND |
| | South Miami, FL | MDWASD South District WWTF | Liquids | 920,528 | 79 | 1/17/23 | 7/27/23 | ND | 18433 | ND |
| | South, Orange County, FL | South Water Reclamation Facility | Solids (Settling w/ Imhoff | 183,009 | 186 | 4/5/22 | 7/27/23 | ND | 107546 | ND |

| | | | | | | | | | | |
|---|---|---|---|---|---|---|---|---|---|---|
| | | | cones onsite) | | | | | | | |
| | Tallahassee, FL | TPSmith Water Reclamation Facility | | 212,065 | 97 | 10/30/22 | 7/27/23 | ND | 307698 | ND |
| | Southwest, Orange County, FL | Hamlin Water Reclamation Facility | Liquids | 50,000 | 5 | 7/16/23 | 7/27/23 | ND | 13548 | ND |
| Georgia | Big Creek, Roswell, GA | Big Creek Water Reclamation Facility | Liquids | 189,593 | 161 | 6/28/22 | 7/24/23 | ND | 206693 | 10379 |
| | College Park, GA | Camp Creek Water Reclamation Facility | Liquids | 73,821 | 164 | 6/29/22 | 7/24/23 | ND | 93666 | 7306 |
| | Columbus, GA | South Columbus Water Resources Facility | Solids | 278,000 | 109 | 8/17/22 | 7/18/23 | ND | 106952 | 4254 |

| | | | | | | | | | | |
|---|---|---|---|---|---|---|---|---|---|---|
| | Johns Creek, Roswell, GA | Johns Creek Environmental Campus | Liquids | 84,486 | 160 | 6/28/22 | 7/24/23 | ND | 219469 | 10365 |
| | Little River, Roswell, GA | Little River Water Reclamation Facility | Liquids | 12,818 | 160 | 6/29/22 | 7/24/23 | ND | 176038 | 4752 |
| | RM Clayton, Atlanta, GA | RM Clayton Water Reclamation Center | Liquids | 294,660 | 108 | 11/1/22 | 7/27/23 | ND | 93901 | ND |
| | South River, Atlanta, GA | South River Water Reclamation Center | Liquids | 105,160 | 107 | 11/1/22 | 7/27/23 | ND | 86826 | ND |
| | Utoy Creek, Atlanta, GA | Utoy Creek Water Reclamation Center | Liquids | 70,887 | 107 | 11/1/22 | 7/27/23 | ND | 94549 | ND |
| Hawaii | Honolulu, HI | Honouliuli Wastewater Treatment Plant | Liquids | 300,000 | 11 | 5/10/23 | 6/2/23 | ND | 500 | ND |

| | | | | | | | | | | |
|---|---|---|---|---|---|---|---|---|---|---|
| | Honolulu, HI | Kailua Regional Wastewater Treatment Plant | Liquids | 90,000 | 11 | 6/28/23 | 7/24/23 | ND | 4822 | ND |
| | Honolulu, HI | Sand Island Wastewater Tratment Plant | Liquids | 390,000 | 11 | 6/28/23 | 7/24/23 | ND | 5450 | ND |
| | Honolulu, HI | Waianae Wastewater Treatment Plant | Liquids | 44,000 | 11 | 6/28/23 | 7/24/23 | ND | 6505 | ND |
| | Honolulu, HI | Wahiawa Wastewater Treatment Plant | Liquids | 18,000 | 11 | 6/28/23 | 7/24/23 | ND | 5071 | ND |
| | Hilo, HI | Hilo Wastewater Treatment Plant | Liquids | 16,257 | 11 | 6/28/23 | 7/24/23 | ND | 6173 | ND |

| | | | | | | | | | | |
|---|---|---|---|---|---|---|---|---|---|---|
| Idaho | Coeur d'Alene, ID | City of Coeur d'Alene Water Resource Recovery Facility | Solids | 50,540 | 201 | 3/7/22 | 7/26/23 | ND | 130638 | ND |
| | Lander Street, Boise, ID | Lander Street Water Renewal Facility | Liquids | 108,556 | 82 | 1/18/23 | 7/26/23 | ND | 129667 | ND |
| | West Boise, ID | West Boise Water Renewal Facility | Liquids | 186,901 | 82 | 1/18/23 | 7/26/23 | ND | 68986 | ND |
| Illinois | Glen Ellyn, IL | Glenbard Wastewater Authority | Solids (Settling w/ Imhoff cones onsite) | 86,000 | 139 | 8/8/22 | 7/25/23 | ND | 82154 | 2889 |
| | Wheaton, IL | Wheaton Sanitary District | Solids (Settling w/ Imhoff cones onsite) | 63,000 | 131 | 9/16/22 | 7/26/23 | ND | 101632 | 5005 |

| | | | | | | | | | | |
|---|---|---|---|---|---|---|---|---|---|---|
| Indiana | Bloomington, IN | Dillman Road WWTP | Liquids | 56,090 | 135 | 8/18/22 | 7/25/23 | ND | 227767 | 6578 |
| | Carmel, IN | City of Carmel WWTP | Solids | 86,000 | 36 | 5/3/23 | 7/26/23 | ND | 3984 | ND |
| | Downtown, Jeffersonville, IN | Jeffersonville Downtown WWTP | Liquids | 25,000 | 111 | 10/28/22 | 7/26/23 | ND | 166791 | 4206 |
| | North, Jeffersonville, IN | North Water Reclamation Facility | Liquids | 25,000 | 109 | 10/28/22 | 7/26/23 | ND | 57764 | ND |
| | South Bend, IN | City of South Bend Wastewater Treatment Plant | Liquids | 130,000 | 121 | 9/13/22 | 7/27/23 | ND | 147468 | 3577 |
| | Clinton, IA | City of Clinton | Solids | 29,300 | 81 | 1/18/23 | 7/26/23 | ND | 3197 | ND |

Iowa

| | | | | | | | | | | |
|---|---|---|---|---|---|---|---|---|---|---|
| | Coralville, IA | Coralville Wastewater Treatment Facility | Liquids | 23,000 | 76 | 1/25/23 | 7/26/23 | ND | 88157 | ND |
| | Marshalltown, IA | City of Marshalltown Water Pollution Control Plant | Liquids | 27,400 | 78 | 1/24/23 | 7/27/23 | ND | 20979 | ND |
| | Muscatine, IA | Muscatine STP | Solids | 24,400 | 95 | 12/14/22 | 7/24/23 | ND | 84422 | ND |
| | Ottumwa, IA | Ottumwa WPCF | Liquids | 25,529 | 94 | 12/19/22 | 7/26/23 | ND | 41198 | ND |
| Kansas | Kaw Point, Kansas City, KS | Municipal Wastewater Treatment Plant No. 1 (Kaw Point) | Solids (Settling w/ Imhoff cones onsite) | 90,000 | 82 | 1/12/23 | 7/25/23 | ND | 13946 | ND |
| | Lawrence, KS | Lawrence Kansas River Wastewater | Solids (Settling w/ Imhoff cones onsite) | 80,000 | 140 | 8/3/22 | 7/26/23 | ND | 76170 | 5339 |

| | | | | | | | | | | |
|---|---|---|---|---|---|---|---|---|---|---|
| | | Treatment Facility | | | | | | | | |
| | P20, Kansas City, KS | Kansas City Treatment Plant #20 | Solids (Settling w/ Imhoff cones onsite) | 35,000 | 83 | 1/12/23 | 7/25/23 | ND | 30173 | ND |
| | Salina, KS | Salina Wastewater Treatment Plant | Solids | 47,000 | 151 | 8/10/22 | 7/26/23 | ND | 74362 | 2031 |
| | Wolcott, Kansas City, KS | Wolcott Wastewater Treatment Facility | Liquids | 15,000 | 82 | 1/11/23 | 7/25/23 | ND | 60852 | ND |
| Kentucky | Louisville, KY | Morris Forman Water Quality Treatment Center | Solids (Settling w/ Imhoff cones onsite) | 423,913 | 69 | 3/10/22 | 7/19/23 | ND | 19025 | ND |

| | | | | | | | | | | |
|---|---|---|---|---|---|---|---|---|---|---|
| Maine | Bangor, ME | City of Bangor Wastewater Treatment Plant | Liquids | 40,000 | 19 | 5/17/23 | 7/24/23 | ND | 8708 | ND |
| | Brunswick, ME | Brunswick Sewer District | Liquids | 10,000 | 89 | 12/1/22 | 7/25/23 | ND | 132003 | ND |
| | Portland, ME | Portland Water District (East End Wastewater Treatment Facility) | Liquids | 65,000 | 102 | 9/5/22 | 7/19/23 | ND | 101124 | 5889 |
| | York, ME | York Sewer District | Liquids | 10,000 | 75 | 1/23/23 | 7/24/23 | ND | 8619 | ND |
| Maryland | Hagerstown, MD | Hagerstown Wastewater Treatment Plant | Liquids | 90,000 | 90 | 12/16/22 | 7/26/23 | ND | 80848 | ND |

| | | | | | | | | | | |
|---|---|---|---|---|---|---|---|---|---|---|
| | Hollywood, MD | Marlay Taylor Water Reclamation Facility | Liquids | 55,000 | 75 | 1/6/23 | 7/26/23 | ND | 28310 | ND |
| Massachusetts | Boston, MA | Deer Island Treatment Plant | Solids | 2,400,000 | 92 | 12/14/22 | 7/26/23 | ND | 30877 | 1812 |
| | Millbury, MA | Upper Blackstone Clean Water | Liquids | 250,000 | 61 | 3/1/23 | 7/27/23 | ND | 20063 | ND |
| Michigan | Ann Arbor, MI | City of Ann Arbor Wastewater Treatment Plant | Liquids | 125,000 | 164 | 6/29/22 | 7/25/23 | ND | 165245 | 4273 |
| | Jackson, MI | Jackson Wastewater Treatment Plant | Solids | 90,000 | 197 | 4/21/22 | 7/27/23 | ND | 71206 | 1772 |
| | Jenison, MI | Grandville Clean | Solids | 75,000 | 98 | 12/12/22 | 7/26/23 | ND | 47783 | ND |

| | | | | | | | | | | |
|---|---|---|---|---|---|---|---|---|---|---|
| | | Water Plant | | | | | | | | |
| | Mt. Pleasant, MI | Mt. Pleasant WRRF | Liquids | 21,690 | 42 | 4/11/23 | 7/26/23 | ND | 22143 | ND |
| | Traverse City, MI | Traverse City Regional Waste Water Treatment Plant | Liquids | 30,623 | 76 | 1/25/23 | 7/24/23 | ND | 37865 | ND |
| | Warren, MI | City of Warren Wastewater Treatment Plant | Liquids | 140,000 | 126 | 9/29/22 | 7/25/23 | ND | 305659 | 3130 |
| Minnesota | Mankato, MN | City of Mankato Water Resource Recovery Facility (WRRF) | Solids (Settling w/ Imhoff cones onsite) | 70,000 | 139 | 8/31/22 | 7/25/23 | ND | 52217 | 2461 |

| | | | | | | | | | | |
|---|---|---|---|---|---|---|---|---|---|---|
| | Red Wing, MN | Red Wing Wastewater Treatment Facility | Solids (Settling w/ Imhoff cones onsite) | 16,000 | 33 | 5/10/23 | 7/26/23 | ND | 2965 | ND |
| | Rochester, MN | City Of Rochester MN Water Reclamation Plant | Solids | 120,000 | 112 | 11/7/22 | 7/26/23 | ND | 85742 | 2372 |
| | St. Cloud, MN | St. Cloud Nutrient, Energy and Water Recovery Facility | Liquids | 120,000 | 49 | 4/5/23 | 7/26/23 | ND | 13536 | ND |
| Nevada | Las Vegas, NV | Clark County Water Reclamation District (CCWRD) Flamingo Water Resource Center (FWRC) | Liquids | 990,000 | 52 | 3/29/23 | 7/26/23 | ND | 23685 | ND |

| | | | | | | | | | | |
|---|---|---|---|---|---|---|---|---|---|---|
| New Hampshire | Dover, NH | City of Dover Wastewater Treatment Facility | Liquids | 30,000 | 93 | 12/1/22 | 7/25/23 | ND | 188575 | ND |
| | Hall Street, Concord, NH | Hall Street Wastewater Treatment Plant | Liquids | 45,000 | 122 | 10/14/22 | 7/26/23 | ND | 158768 | ND |
| | Penacook, Concord, NH | Penacook Wastewater Treatment Facility | Liquids | 4,000 | 120 | 10/14/22 | 7/26/23 | ND | 257215 | ND |
| New Jersey | Belmar, NJ | South Monmouth Regional Sewerage Authority | Solids | 52,672 | 99 | 12/7/22 | 7/26/23 | ND | 73594 | 2336 |
| | Bridgeton, NJ | Cumberland County Utilities Authority | Liquids | 50,000 | 54 | 3/15/23 | 7/26/23 | ND | 3986 | ND |

| | | | | | | | | | | |
|---|---|---|---|---|---|---|---|---|---|---|
| | Bridgewater, NJ | The Somerset Raritan Valley Sewerage Authority | Liquids | 130,000 | 29 | 5/17/23 | 7/26/23 | ND | ND | ND |
| | Newark, NJ | Passaic Valley Sewerage Commission | Solids | 1,500,000 | 135 | 8/8/22 | 7/25/23 | ND | 84335 | 3072 |
| | Oakhurst, NJ | Township of Ocean Sewerage Authority | Liquids | 50,000 | 29 | 4/13/23 | 7/25/23 | ND | 60801 | ND |
| | Union Beach, NJ | Bayshore Regional Sewerage Authority | Solids | 100,000 | 38 | 5/1/23 | 7/26/23 | ND | 2889 | ND |
| North Carolina | Kinston, NC | Johnnie Mosley Regional Water Reclamation Facility | Liquids | 25,000 | 95 | 10/19/22 | 7/26/23 | ND | 88909 | 1852 |

| | | | | | | | | | | |
|---|---|---|---|---|---|---|---|---|---|---|
| | Winston-Salem, NC | Archie Elledge WWTP | Liquids | 92,000 | 130 | 8/24/22 | 7/26/23 | ND | 120049 | ND |
| Ohio | Akron, OH | Akron Water Reclamation Facility | Solids | 365,000 | 77 | 1/9/23 | 7/26/23 | ND | 16366 | ND |
| | Youngstown, OH | City of Youngstown Wastewater Treatment Plant | Solids | 174,000 | 93 | 12/16/22 | 7/26/23 | ND | 39825 | ND |
| Pennsylvania | Chester, PA | DELCORA Western Regional Treatment Plant | Liquids | 220,000 | 101 | 11/1/22 | 7/27/23 | ND | 62722 | ND |
| | Harrisburg, PA | Capital Region Water AWTF | Liquids | 125,000 | 154 | 8/4/22 | 7/27/23 | ND | 110394 | 3318 |

| | | | | | | | | | | |
|---|---|---|---|---|---|---|---|---|---|---|
| South Dakota | Yankton, SD | City of Yankton Wastewater Treatment Facility | Liquids | 20,000 | 38 | 4/27/23 | 7/27/23 | ND | ND | ND |
| Tennessee | Chattanooga, TN | Moccasin Bend Wastewatet Treatment Plant | Liquids | 400,000 | 18 | 6/14/23 | 7/26/23 | ND | 3111 | ND |
| Texas | Gainesville, TX | City of Gainesville Wastewater Treatment Plant | Liquids | 17,300 | 8 | 7/3/23 | 7/24/23 | ND | 2700 | ND |
| | Dallas, TX | DCWT Dallas | Liquids | 270,000 | 81 | 12/27/22 | 7/26/23 | ND | 904894 | ND |
| | Dallas, TX | DCWT White Workc | Liquids | 630,000 | 212 | 3/10/22 | 7/26/23 | ND | 70946 | 3564 |
| | Dallas, TX | Southside Wastewater | Liquids | 421,700 | 96 | 12/12/22 | 7/25/23 | ND | 143227 | ND |

| | | | | | | | | | | |
|---|---|---|---|---|---|---|---|---|---|---|
| | | Treatment Plant | | | | | | | | |
| | Garland, TX | City of Garland Rowlett Creek WWTP | Solids | 200,000 | 92 | 12/6/22 | 7/25/23 | ND | 138798 | ND |
| | Hollywood Road, Amarillo, TX | Hollywood Road WWTP | Liquids | 60,000 | 84 | 12/14/22 | 7/26/23 | ND | 103746 | ND |
| | River Road, Amarillo, TX | River Road WWTP | Liquids | 140,000 | 9 | 6/25/23 | 7/13/23 | ND | 4721 | ND |
| | South, Laredo, TX | South Laredo WWTP | Liquids | 120,000 | 169 | 3/1/22 | 5/1/23 | ND | 25341 | ND |
| | Sunnyvale, TX | Duck Creek Wastewate r Treatment Plant | Solids | 186,000 | 10 | 7/3/23 | 7/24/23 | ND | ND | ND |

| | | | | | | | | | | |
|---|---|---|---|---|---|---|---|---|---|---|
| | Wichita Falls, TX | Wichita Falls Resource Recovery Facility | Solids | 90,000 | 97 | 12/7/22 | 7/26/23 | ND | 42563 | ND |
| | Woodlands SJRA WWTF No. 1, TX | SJRA WWTF No.1 | Liquids | 65,000 | 67 | 2/22/23 | 7/26/23 | ND | 18409 | ND |
| | Woodlands SJRA WWTF No. 2, TX | SJRA WWTF No.2 | Liquids | 70,000 | 67 | 2/22/23 | 7/26/23 | ND | 22538 | ND |
| | Woodlands SJRA WWTF No. 3, TX | SJRA WWTF No.3 | Liquids | 15,000 | 67 | 2/22/23 | 7/26/23 | ND | 34219 | ND |
| Utah | Zacate Creek, Laredo, TX | Zacate Creek WWTP | Liquids | 140,000 | 82 | 12/14/22 | 7/26/23 | ND | 87144 | ND |
| | Central Salt Lake Valley, UT | Central Valley Water Reclamation Facility | Solids | 600,000 | 114 | 11/2/22 | 7/25/23 | ND | 153973 | 3393 |

| | | | | | | | | | | |
|---|---|---|---|---|---|---|---|---|---|---|
| Vermont | Provo, UT | Provo City Water Reclamation Facility | Solids (Settling w/ Imhoff cones onsite) | 115,000 | 87 | 9/21/22 | 7/24/23 | ND | 158712 | 3042 |
| | Essex Junction, VT | City of Essex Junction Wastewater Treatment Facility | Solids | 30,000 | 56 | 3/6/23 | 7/26/23 | ND | 34147 | ND |
| | Montpelier, VT | Montpelier Water Resource Recovery Facility | Solids | 10,100 | 55 | 3/9/23 | 7/20/23 | ND | 31115 | ND |
| Virginia | South Burlington, VT | South Burlington-Airport Parkway WWTF | Liquids | 16,000 | 32 | 3/28/23 | 7/25/23 | ND | ND | ND |
| | Aquia, Stafford, VA | Aquia Wastewater Treatment Facility | Solids | 100,000 | 63 | 3/3/23 | 7/26/23 | ND | ND | ND |

| | | | | | | | | | | |
|---|---|---|---|---|---|---|---|---|---|---|
| | Hillsville, VA | Town of Hillsville Wastewater Treatment Plant | Solids | 3,000 | 96 | 12/14/22 | 7/26/23 | ND | 3407 | ND |
| West Virginia | Little Falls Run, Stafford, VA | Little Falls Run Wastewater Treatment Facility | Solids | 50,000 | 62 | 3/3/23 | 7/26/23 | ND | 2254 | ND |
| | Wheeling, WV | City of Wheeling, Water Pollution Control Division | Solids | 100,000 | 44 | 4/12/23 | 7/24/23 | ND | ND | ND |
| Wisconsin | Wausau, WI | Wausau Waterworks Treatment Facility | Liquids | 44,000 | 11 | 6/20/23 | 7/27/23 | ND | ND | ND |

Table S2. Forward and reverse primers, and probe sequences for detection of viral nucleic acids in this study. Primers and probes were purchased from Integrated DNA Technologies (IDT, Coralville, Iowa). All probes contained fluorescent molecules (see Figure S1) and quenchers (ZEN/3′ IBFQ). ZEN, a proprietary internal quencher from IDT; IBFQ, Iowa Black FQ. See Boehm et al.[3] for more details of the assays.

| Target | Primer/Probe | Sequence |
|---|---|---|
| BCoV | Forward | CTGGAAGTTGGTGGAGTT |
| | Reverse | ATTATCGGCCTAACATACATC |
| | Probe | CCTTCATATCTATACACATCAAGTTGTT |
| PMMoV | Forward | GAGTGGTTTGACCTTAACGTTTGA |
| | Reverse | TTGTCGGTTGCAATGCAAGT |
| | Probe | CCTACCGAAGCAAATG |
| RSV | Forward | CTCCAGAATAYAGGCATGAYTCTCC |
| | Reverse | GCYCTYCTAATYACWGCTGTAAGAC |
| | Probe | TAACCAAATTAGCAGCAGGAGATAGATCAG |

**Figure S1.** A schematic of how assays were multiplexed during different periods of the project.

Row A is for all WWTPS in the study except for Gilroy, CA; Oceanside San Francisco, CA; Palo Alto, CA; Redwood City, CA; Sacramento, CA; San Jose, CA; Southeast San Francisco, CA; and Sunnyvale, CA. Row B is for Gilroy, CA; Oceanside San Francisco, CA; Palo Alto, CA; Redwood City, CA; Sacramento, CA; San Jose, CA; Southeast San Francisco, CA; and Sunnyvale, CA. Note that samples from Modesto and Merced, CA were processed with reactions in row B until 1 May 2022 after which they were processed with reactions in Row A. The date at the bottom left side of each box is the start date and the date near the right edge of the box is the approximate end date (based on the date the assay was stopped in the lab, the date associated with the last sample run could be different depending on the date it was processed in the lab). All probes contained fluorescent molecules as indicated in parentheses: FAM, 6-fluorescein amidite; HEX, hexachloro-fluorescein, Cy5, Cyanine-5; Cy5.5, Cyanine5.5; ROX, carboxyrhodamine; and ATTO59. If the box is white, the annealing temperature is 59 °C and if gray, it is 61 °C. N and S are assays targeting those genes in SARS-CoV-2. HV69-70 is the assay targeting the deletion 69/70 in the S gene characteristic of Alpha, and various Omicron variants. BA.2.75 is the assay targeting the adjunct SNPs in the S gene characteristic of BA.2.75. RSV is the assay targeting a gene in the RSV genome. BA.2 is the assay targeting the set of deletions LPPA24S characteristic of BA.2. IBV is the assay targeting influenza B. IAV is the assay targeting influenza A. XBB* is an assay targeted adjacent SNPs in the SARS-CoV-2 XBB* sublineages. Noro G2 is an assay targeting the genome of human norovirus GII. MPXV_G2R_G is the assay targeting MPXV clade II.

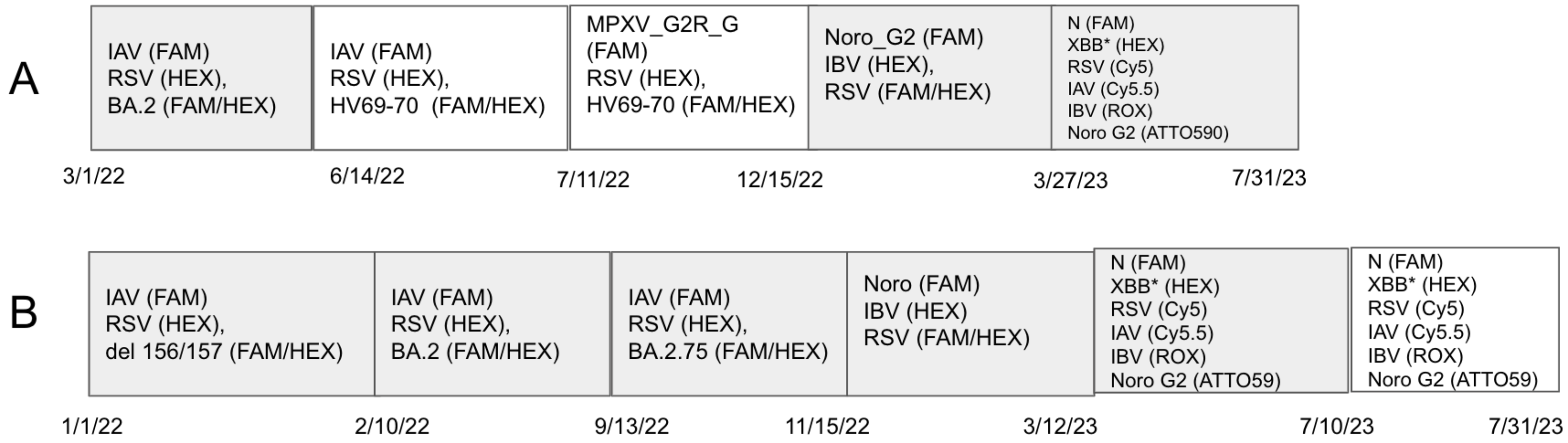

**Figure S2.** RSV concentrations measured at each WWTP included in this study. The errors on each data point represent standard deviations.

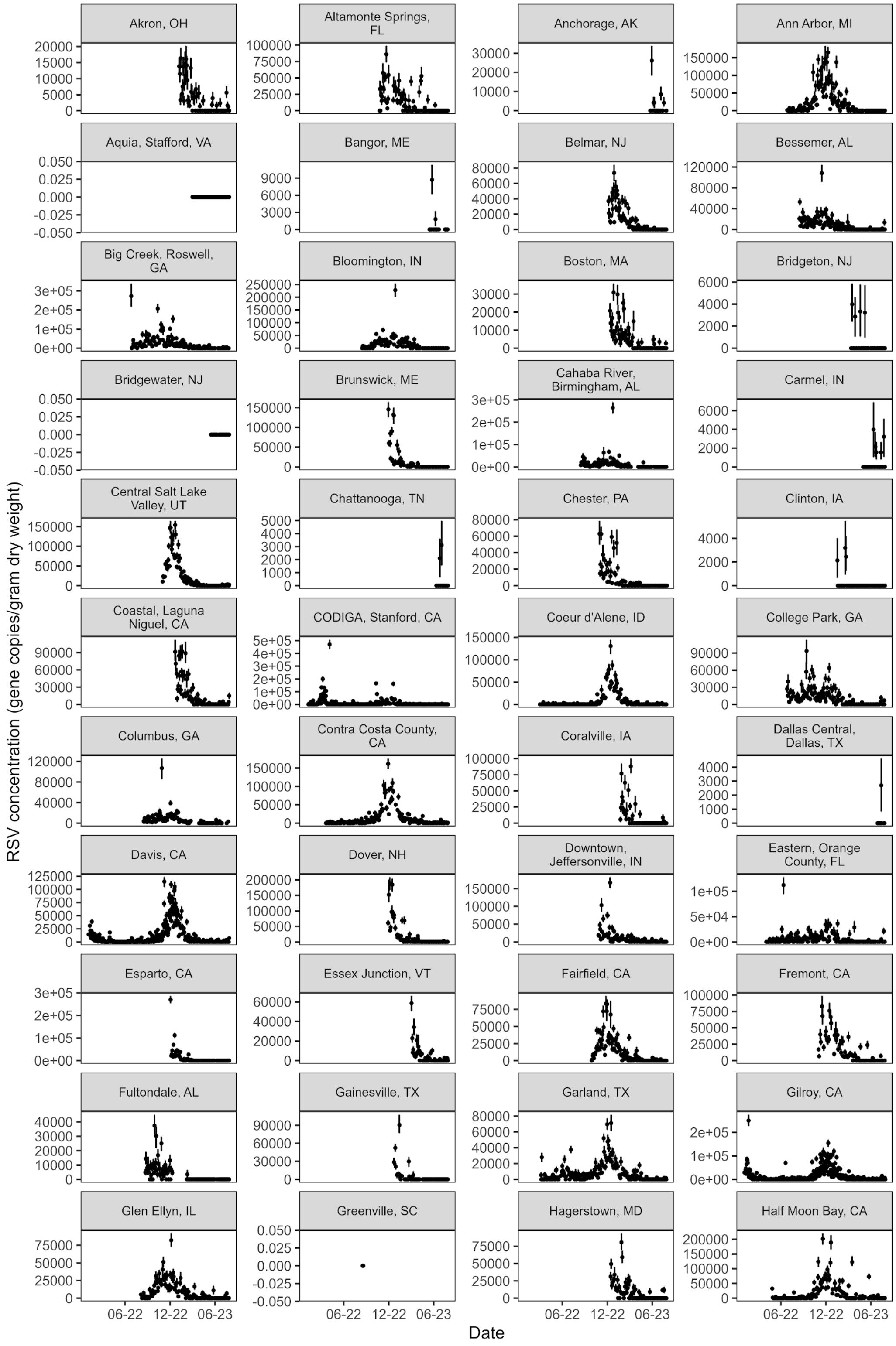
Akron, OH
Altamonte Springs, FL
Anchorage, AK
Ann Arbor, MI
Aquia, Stafford, VA
Bangor, ME
Belmar, NJ
Bessemer, AL
Big Creek, Roswell, GA
Bloomington, IN
Boston, MA
Bridgeton, NJ
Bridgewater, NJ
Brunswick, ME
Cahaba River, Birmingham, AL
Carmel, IN
Central Salt Lake Valley, UT
Chattanooga, TN
Chester, PA
Clinton, IA
Coastal, Laguna Niguel, CA
CODIGA, Stanford, CA
Coeur d'Alene, ID
College Park, GA
Columbus, GA
Contra Costa County, CA
Coralville, IA
Dallas Central, Dallas, TX
Davis, CA
Dover, NH
Downtown, Jeffersonville, IN
Eastern, Orange County, FL
Esparto, CA
Essex Junction, VT
Fairfield, CA
Fremont, CA
Fultondale, AL
Gainesville, TX
Garland, TX
Gilroy, CA
Glen Ellyn, IL
Greenville, SC
Hagerstown, MD
Half Moon Bay, CA
06-22
12-22
06-23
Date
RSV concentration (gene copies/gram dry weight)

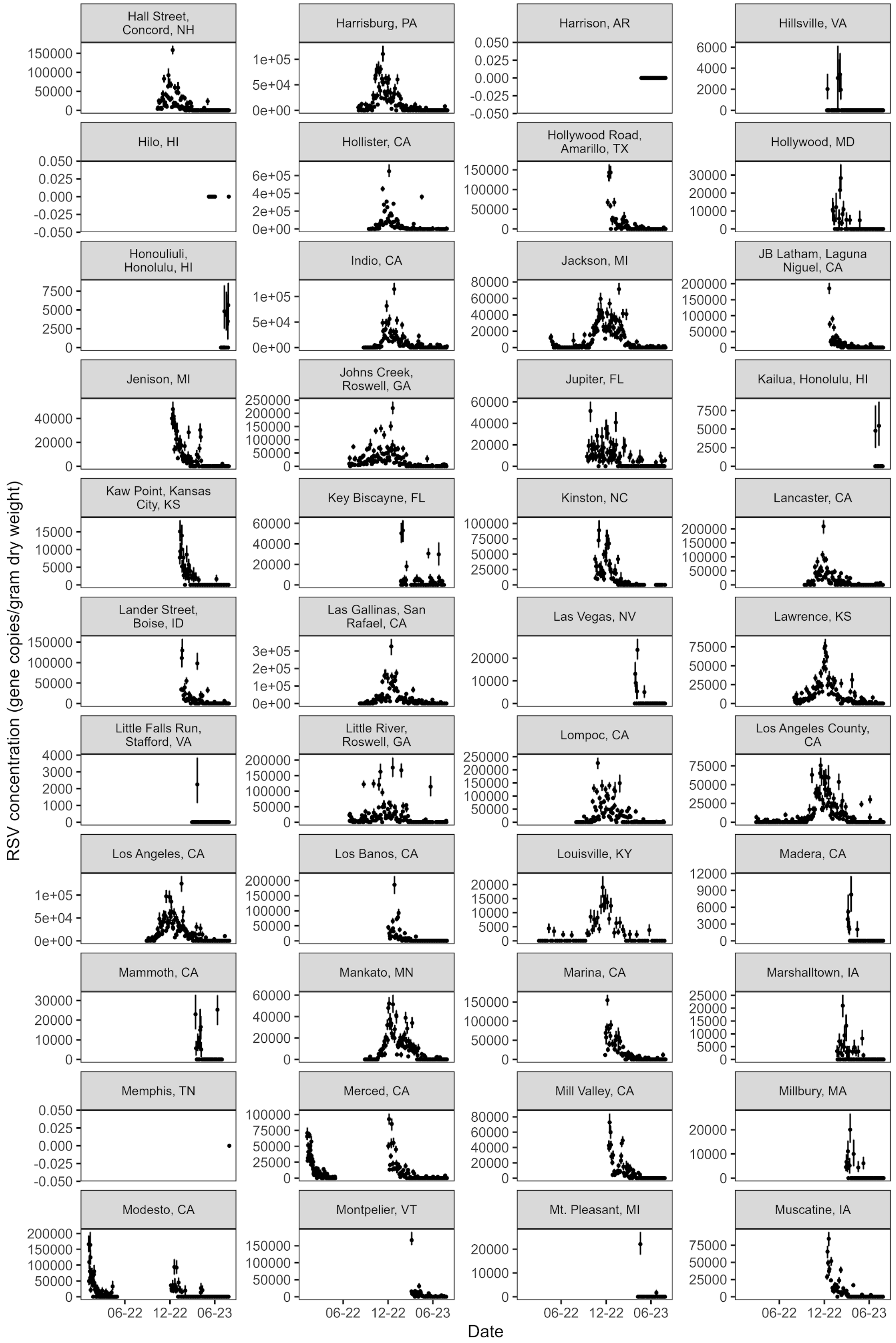

Hall Street, Concord, NH
Harrisburg, PA
Harrison, AR
Hillsville, VA
Hilo, HI
Hollister, CA
Hollywood Road, Amarillo, TX
Hollywood, MD
Honouliuli, Honolulu, HI
Indio, CA
Jackson, MI
JB Latham, Laguna Niguel, CA
Jenison, MI
Johns Creek, Roswell, GA
Jupiter, FL
Kailua, Honolulu, HI
Kaw Point, Kansas City, KS
Key Biscayne, FL
Kinston, NC
Lancaster, CA
Lander Street, Boise, ID
Las Gallinas, San Rafael, CA
Las Vegas, NV
Lawrence, KS
Little Falls Run, Stafford, VA
Little River, Roswell, GA
Lompoc, CA
Los Angeles County, CA
Los Angeles, CA
Los Banos, CA
Louisville, KY
Madera, CA
Mammoth, CA
Mankato, MN
Marina, CA
Marshalltown, IA
Memphis, TN
Merced, CA
Mill Valley, CA
Millbury, MA
Modesto, CA
Montpelier, VT
Mt. Pleasant, MI
Muscatine, IA
RSV concentration (gene copies/gram dry weight)
06-22
12-22
06-23
Date

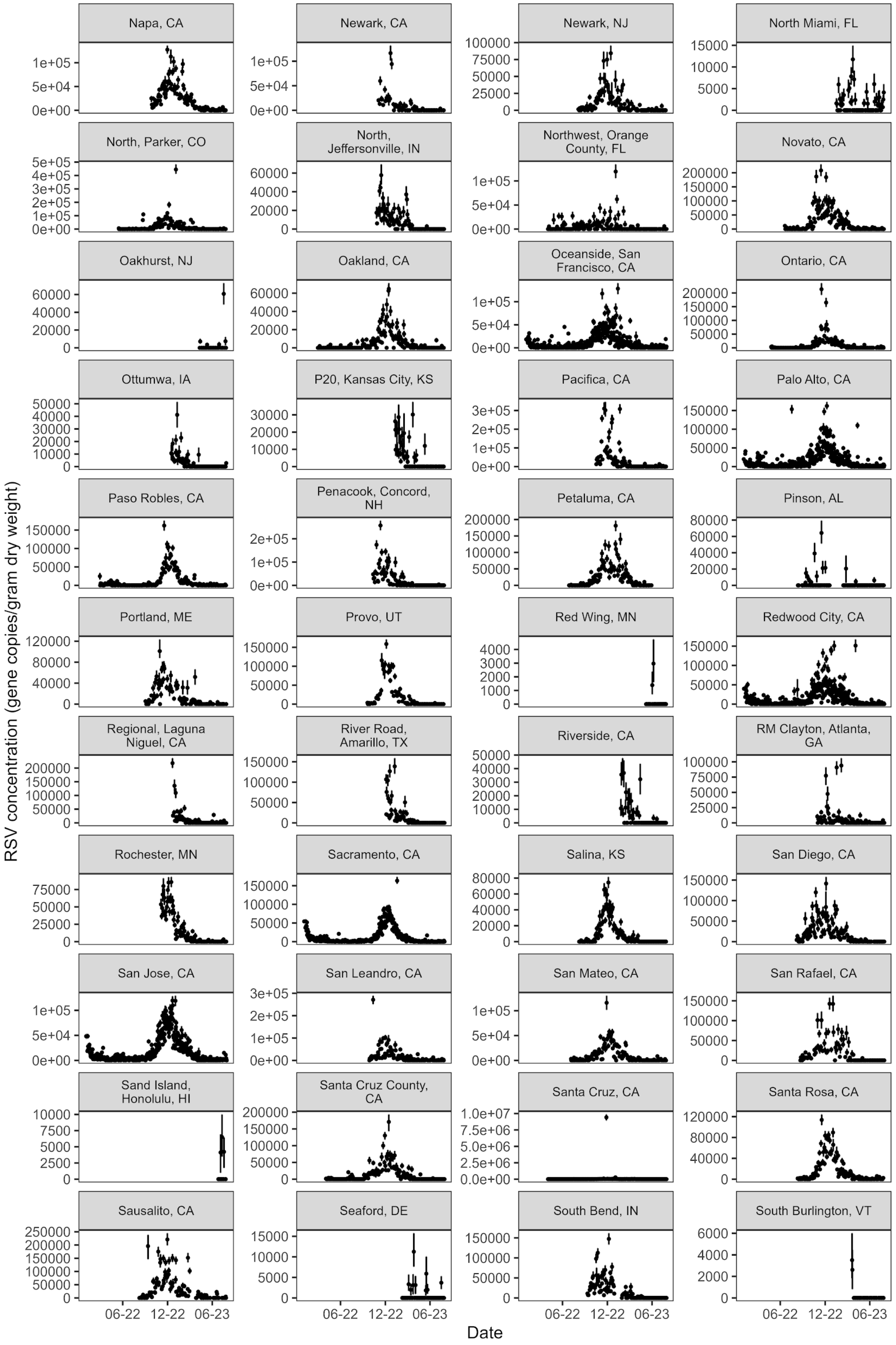
Napa, CA
Newark, CA
Newark, NJ
North Miami, FL
North, Parker, CO
North, Jeffersonville, IN
Northwest, Orange County, FL
Novato, CA
Oakhurst, NJ
Oakland, CA
Oceanside, San Francisco, CA
Ontario, CA
Ottumwa, IA
P20, Kansas City, KS
Pacifica, CA
Palo Alto, CA
Paso Robles, CA
Penacook, Concord, NH
Petaluma, CA
Pinson, AL
Portland, ME
Provo, UT
Red Wing, MN
Redwood City, CA
Regional, Laguna Niguel, CA
River Road, Amarillo, TX
Riverside, CA
RM Clayton, Atlanta, GA
Rochester, MN
Sacramento, CA
Salina, KS
San Diego, CA
San Jose, CA
San Leandro, CA
San Mateo, CA
San Rafael, CA
Sand Island, Honolulu, HI
Santa Cruz County, CA
Santa Cruz, CA
Santa Rosa, CA
Sausalito, CA
Seaford, DE
South Bend, IN
South Burlington, VT
06-22
12-22
06-23
Date
RSV concentration (gene copies/gram dry weight)

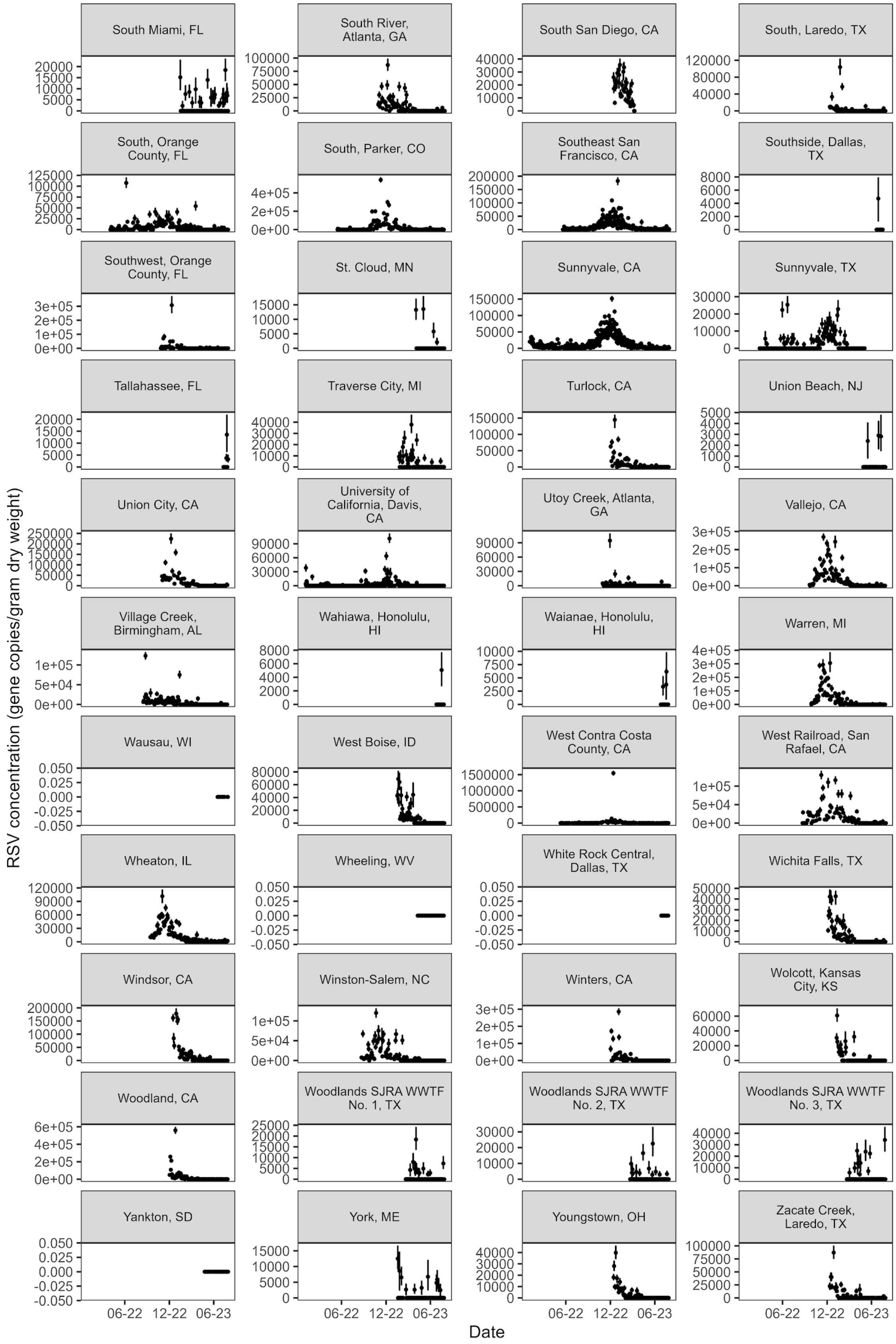
South Miami, FL
South River, Atlanta, GA
South San Diego, CA
South, Laredo, TX
South, Orange County, FL
South, Parker, CO
Southeast San Francisco, CA
Southside, Dallas, TX
Southwest, Orange County, FL
St. Cloud, MN
Sunnyvale, CA
Sunnyvale, TX
Tallahassee, FL
Traverse City, MI
Turlock, CA
Union Beach, NJ
Union City, CA
University of California, Davis, CA
Utoy Creek, Atlanta, GA
Vallejo, CA
Village Creek, Birmingham, AL
Wahiawa, Honolulu, HI
Waianae, Honolulu, HI
Warren, MI
Wausau, WI
West Boise, ID
West Contra Costa County, CA
West Railroad, San Rafael, CA
Wheaton, IL
Wheeling, WV
White Rock Central, Dallas, TX
Wichita Falls, TX
Windsor, CA
Winston-Salem, NC
Winters, CA
Wolcott, Kansas City, KS
Woodland, CA
Woodlands SJRA WWTF No. 1, TX
Woodlands SJRA WWTF No. 2, TX
Woodlands SJRA WWTF No. 3, TX
Yankton, SD
York, ME
Youngstown, OH
Zacate Creek, Laredo, TX
RSV concentration (gene copies/gram dry weight)
06-22
12-22
06-23
Date

**Figure S3.** Availability of wastewater data by state in 2022-2023

Horizontal blue bars represent duration of RSV epidemics in 2022-2023 by state as determined based on NREVSS data (clinical onset dates). Dashed red lines indicate the start and end of the 2022-2023 RSV surveillance year as determined by the CDC. Black dots represent the start of wastewater data collection in each state. States shown in panel A (N=14) had wastewater data available prior to the beginning of the 2022-2023 RSV epidemic and were included in the analysis. States in panel B (N=20) only began wastewater data collection after the onset of the 2022-2023 RSV epidemic and were excluded from the onset and peak analysis (called "analysis in the figure").

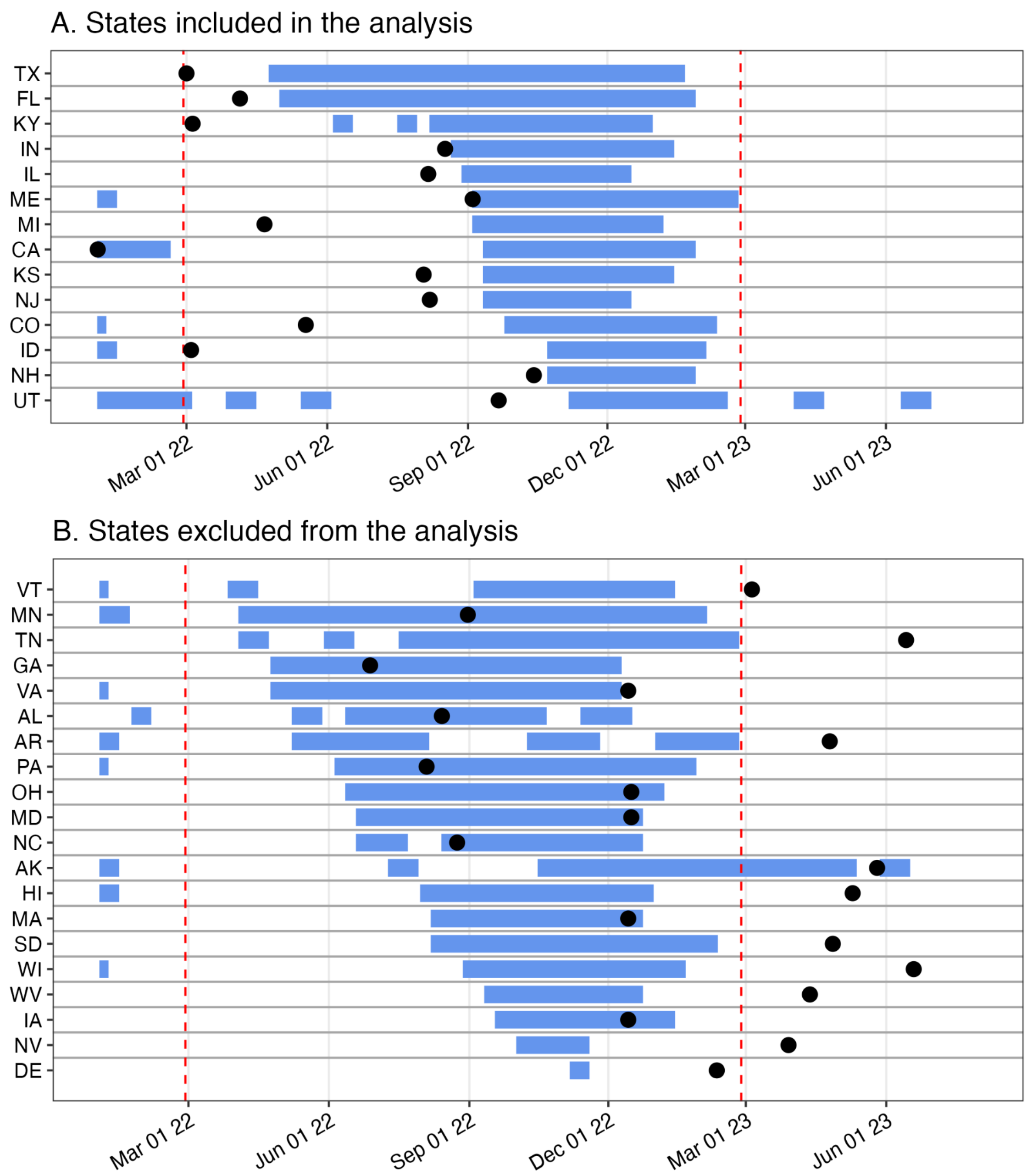

**Figure S4.** Map of wastewater and clinical laboratory surveillance sites for 2022-2023 surveillance year.

Wastewater treatment plants participating in the project and clinical laboratories participating in NREVSS are mapped according to counties in which facilities are located. Orange points depict counties in which only wastewater (WW) surveillance is conducted. Blue points show counties with laboratories that are participating in NREVSS, although they may not submit data year round or during the 2022-2023 season. Red points mark counties that have both laboratories participating in NREVSS and wastewater treatment plants participating in this study. States in dark gray (N=14) were included in the onset and peak analysis of this study.

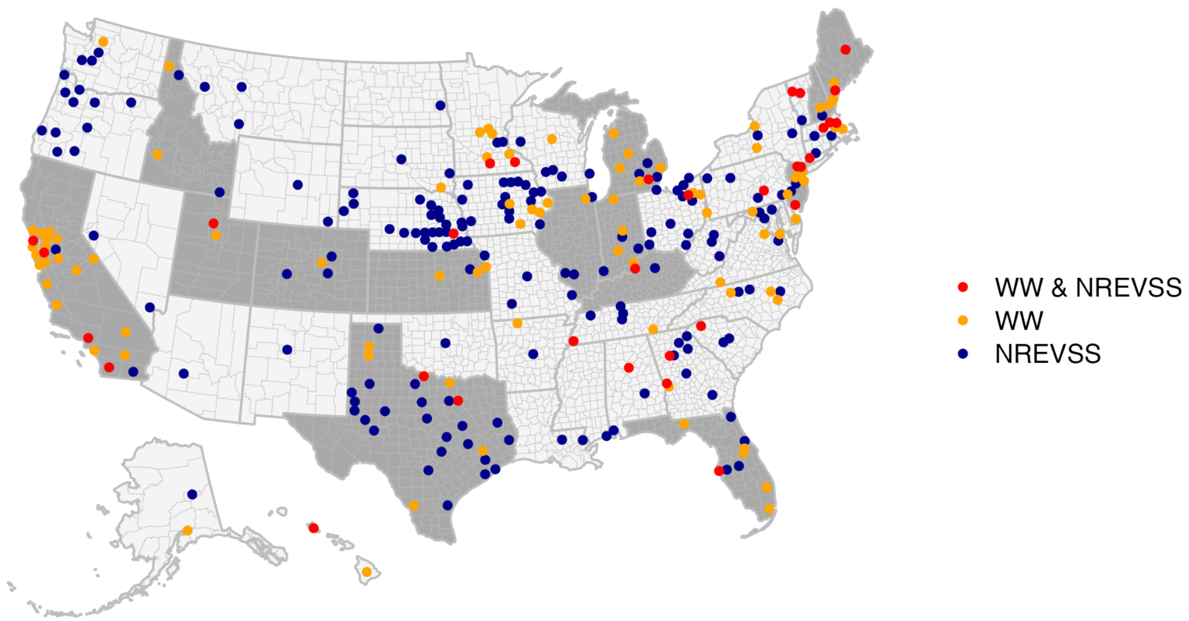
